# Potential spillover effects on diagnostic delay for cancer during the NHS-Galleri trial: a quasi-experimental difference-in-differences study

**DOI:** 10.1101/2024.11.06.24316784

**Authors:** Sean Mann, Pedro Nascimento de Lima, Joshua Eagan, Agne Ulyte, Beth Ann Griffin

**Author notes:** Correspondence to: S Mann.

## Abstract

**Objective:** To examine whether regions participating in the NHS-Galleri trial of a cell-free DNA-based multi-cancer early detection (MCED) test experienced changes in cancer diagnostic delay rates that could indicate the presence of spillover due to constraints on care delivery.

**Design:** Quasi-experimental difference-in-differences analysis.

**Setting:** England, April 2021-September 2024.

**Participants:** All 21 cancer alliance regions in England, 8 of which participated in the trial.

**Main outcomes:** Cancer diagnostic delay rates (primary outcome) and referral rates (secondary outcome). Primary analysis focused on the group of three cancer types (head and neck, lung, upper gastrointestinal) that are prespecified in the trial protocol, have higher signal detection for cell-free DNA-based testing, and were not subject to routine screening. A difference-in-differences analysis was used to compare outcomes in regions participating in NHS-Galleri to regions that did not participate, before and after the start of the trial.

**Results:** Diagnostic delay rates increased in the first six months of the trial for the primary group of three cancer types in participating regions compared to non-participating regions (adjusted difference-in-differences 3.4 percentage points, 95% CI: 1.9-5.0, p<0.001). These differences persisted from 6 to 12 months after trial start and receded over the following two years. No significant change in referral rates for this group of cancer types was observed.

**Conclusions:** Participation in the NHS-Galleri trial was associated with increased diagnostic delay rates for the primary group of high-detection cancer types. The trial may have led to higher demand for limited diagnostic resources (e.g. imaging, endoscopy) required for follow-up of suspected cancer arising from positive MCED test results or usual care processes. If patients in the control group faced delayed diagnosis as a result, this would represent spillover during the first year of the trial and could lead to overestimation of MCED benefit.

**Summary Box:** *What is already known on this topic:*

- The NHS-Galleri trial randomized over 140,000 adults to receive usual cancer screening or usual screening plus a new cell-free DNA-based multi-cancer early detection test in 8 out of 21 regions in England.
- During the trial, higher use of scarce cancer diagnostic services by the intervention group could have led to reduced availability of services for members of the control group and patients not enrolled in the trial.
- The potential for this form of spillover to bias trial results and affect patient access to care is not addressed in the medical literature, guidance on clinical trial methods, or the NHS-Galleri trial protocol.

*What this study adds:*

- In the first year of the trial, participating regions experienced a modest increase in region-wide diagnostic delay rates for cancer types having higher signal detection for cell-free DNA-based testing.
- Increased diagnostic delays could contribute to stage shift for some control group patients and lead to overestimation of the Galleri test’s effect on early detection of cancer.
- Future trials of multi-cancer early detection tests and other interventions that affect utilization of limited healthcare resources should address the potential for spillover due to constraints on care delivery to affect patient outcomes and trial results.

## Introduction

Detecting treatable cancer early is a public health priority. Randomized controlled trials conducted in real-world health settings are used to determine whether new screening tests,^1^ patient navigation services,^2,3^ or automated reminder systems^4,5^ lead to earlier detection of cancer. Interventions that prove effective in these trials are often integrated into clinical practice guidelines and adopted broadly across health systems.^6–8^ Some trials in real-world settings may suffer from an overlooked source of bias, however. Diagnostic resources such as specialist appointments, imaging equipment, or laboratory staff are often limited.^9–11^ During a trial, an intervention that increases utilization of a scarce resource by some patients can reduce availability of that resource for other patients. This can negatively affect outcomes for patients in the control group and lead trial investigators to overestimate intervention benefit.^12^ Economists refer to this problem as “negative spillover” or “crowding out” and randomized evaluations of public health interventions have been designed to avoid or test for its presence.^13–15^ However, this threat to study validity is overlooked in the medical literature and is not addressed in guidance on clinical trial design,^16,17^ reporting,^18–20^ or risk of bias assessment.^21–23^

The ongoing NHS-Galleri trial is a randomized trial in which spillover due to constraints on care delivery may be a concern. This trial invited approximately 1.5 million patients to participate and subsequently randomized over 140,000 adults (50-77 years) to either receive usual care (“standard-of-care and existing screening modalities”) or usual care plus annual screening using a new multi-cancer early detection (MCED) screening test (Galleri^®^).^1^ The Galleri test relies on cell-free DNA (cfDNA) and artificial intelligence to screen for more than 50 different types of cancer.^24^ Participants with suspected cancer—whether identified via usual care or a positive Galleri test—were referred for follow-up assessment to confirm or rule out a cancer diagnosis.^1^ NHS capacity shortfalls and long wait times for cancer diagnosis have been a long-standing concern,^25^ however, and record high referral numbers and rates of delayed follow-up appointments were reported in 2021 as the NHS-Galleri trial was starting.^26^ Additional patient referrals due to Galleri test rollout may have increased strain on diagnostic services and led to higher rates of delayed diagnosis, including for patients in the control arm of the trial. This could lead to biased estimation of the Galleri test’s ability to achieve the trial’s primary objective: reduced incidence of late stage cancer at time of diagnosis.^1^ If this type of spillover occurred, Galleri MCED screening might underperform expectations if adopted broadly across the NHS.

We use provider-level NHS data on cancer diagnostic delays to examine whether spillover occurred during the NHS-Galleri trial. We focus our analyses on the group of three cancer types (lung, head & neck, upper gastrointestinal) that the Galleri test developers identified as “having higher cancer signal detection” for cfDNA-based MCED testing and which were not already subject to routine single-cancer screening.^27^ We hypothesized that, once the trial started, participating regions would experience a change in diagnostic delays when compared to regions not participating in the trial. Such a change could suggest that spillover, in the form of delayed diagnosis for patients in the control group, occurred during the trial. This is an independent analysis as no study contributors are involved in the conduct of the NHS-Galleri trial.

## Methods

### Data sources and study population

We obtained monthly provider-level data on cancer diagnostic delays and staffing from three publicly available NHS England datasets. The NHS England Provider-based Cancer Waiting Times dataset reports the number of referrals from each health provider organization that lead to diagnostic resolution in a given month.^28^ Within these data, referrals are categorized into groups based on time elapsed between referral and diagnostic resolution: 0–14 days, 15–28 days, 29–42 days, 43–62 days, and more than 62 days. We excluded referrals marked as missing or invalid, for non-cancer symptoms, or not associated with a specific tumor site. Remaining referral data were reported separately for 14 suspected cancer types. These data do not distinguish between referrals that result in a cancer diagnosis and those that rule cancer out; both forms of diagnostic resolution are included in our analysis.

We used provider codes to link the cancer waiting times dataset to two other NHS datasets containing healthcare staff numbers,^29^ absences, and the number of hospital beds occupied by confirmed COVID-19 patients.^30^ Linked data were aggregated up to the level of cancer alliance regions using NHS provider, integrated care board, and cancer alliance linking files.^31,32^ Providers not in linking files were manually linked to regions following review of provider websites and locations (appendix table 1). We excluded providers not in the waiting times dataset and providers serving multiple regions.

The resulting dataset contained monthly counts of total referrals and referrals that breached the 28-day faster diagnosis standard in each region. All 21 cancer alliance regions in England were included in our analysis, as were all months for which data were available (as of July 22, 2025), spanning the period from April 2021 through September 2024. This study period includes a six-month pre-trial period and the first three years of the trial. Data were not available prior to April 2021, limiting the number of data points in the pre-trial period.

### Exposure to the NHS-Galleri trial

The NHS-Galleri trial is being conducted in 8 of the 21 cancer alliance regions in England (appendix fig 1). Trial investigators selected participating regions based on factors that included ethnic diversity, socioeconomic status, and relatively higher rates of late-stage cancer diagnosis and cancer mortality.^1^

Trial enrollment began August 31, 2021, with participants providing blood samples for the Galleri test at time of enrollment.^1^ Enrollment was underway in all eight participating regions by mid-November 2021,^33^ though exact start dates by region were unavailable. Enrollment continued through July 16, 2022.^1^ The intervention group received three Galleri screening tests provided on an annual basis, delivered throughout the year: one at enrollment, another twelve months later, and a final test 24 months following enrollment. The final round of blood draws for MCED testing ended in July 2024.^34^

Patients assigned to the intervention group who received positive Galleri test results were referred within 30 days after blood draw for follow-up diagnostic testing.^35^ Following referral—whether due to a positive MCED test, standard single-cancer screening, or symptomatic presentation in usual care—patients proceeded through a standard diagnostic pathway for suspected cancer.^1,36^

Given this timeline, we treated October 2021 as the first month in which participating regions were exposed to the NHS-Galleri trial. This specification relies on the assumption that Galleri testing, referral in cases of positive results, and diagnostic investigation generally took at least one month following blood draw at enrollment. To accommodate uncertainty in timing of exposure across all eight participating regions we also conducted a sensitivity analysis excluding data from September, October, and November 2021 as a phase-in period. Exposure to the trial continued through the end of the study period in September 2024.

### Outcomes

Our primary outcome measure of interest was the monthly rate of cancer diagnostic delays, aggregated up to the regional level. We followed the NHS faster diagnosis standard in defining rate of cancer diagnostic delay as the percentage of patient referrals for suspected cancer that took longer than 28 days to reach diagnostic resolution.^37^ NHS providers are asked to ensure that no more than 25% of patients experience delays according to this standard.^38^ Our secondary outcome measure was the monthly rate of cancer diagnostic referrals (per 100,000 population) in each region.

### Stratifying Analysis of Outcomes by Cancer Type

The trial protocol focuses on a prespecified list of cancers^1^ that were identified by the Galleri test developers as “having higher cancer signal detection, consistent with their ability to release higher amounts of cfDNA into circulation.”^27^ We expect that any spillover effects on diagnostic delay from Galleri test rollout would be concentrated among these cancers and particularly those not already subject to routine single-cancer screening.

We reviewed suspected cancer types in the NHS England wait times dataset according to whether they contained high-detection cancers from the trial protocol and whether they were subject to routine screening (appendix table 2). Based on this mapping, we stratified our analyses according to three groups of cancer types:

- A primary high-detection group of three cancer types (head & neck, lung, upper gastrointestinal) that exclusively contain high-detection cancers from the trial protocol and were not subject to routine screening.
- A secondary expanded high-detection group of seven cancer types (head & neck, lung, upper gastrointestinal, gynaecological, haematological, lower gastrointestinal, urological) that contain high-detection cancers even if they also contain other cancers or were subject to routine screening.
- A secondary low-detection group of seven cancer types (skin, breast, sarcoma, brain/central nervous system, testicular, acute leukaemia, and cancers of ‘other’ suspected origin) that contain no high-detection cancers.

We also conducted exploratory analyses for each individual cancer type in our dataset.

### Covariates

We included the percentage of absent healthcare staff as a covariate to control for time-varying differences in staffing across regions during the COVID-19 pandemic. This covariate was calculated using monthly data on the number of full-time equivalent NHS staff and the number of staff absent from work due to sickness or self-isolation.^29,30^

### Statistical analysis

We analyzed changes in the rate of cancer diagnostic delay and referral rates in regions that participated in the trial compared to regions that did not participate using a difference-in-differences framework with an event study design. These changes were estimated using a set of variables that captured the interaction between each monthly time period and participation in the NHS-Galleri trial. Coefficient estimates for these variables represent the mean differential change in diagnostic delay or referral rates between participating and non-participating regions for each month during the study period. Months were labeled numerically in relation to the start of the trial in October 2021, which was labeled month 0. The first month for which data were available (April 2021, month −6) served as the reference period. Regression models included month and region fixed effects to control for time invariant differences between region as well as England-wide changes in diagnostic delays over time. Regressions were weighted by the monthly volume of cancer diagnostic referrals in each region (for delay rate analyses) and by population (for referral rate analyses). All models included clustered standard errors at the regional level. Simulation models were used to test robustness of this study design for our specific time series data relative to other candidate methods, finding clear evidence of acceptable power and type I error rates to support use of this differences-in-differences framework (appendix methods 1).

We used the same regression approach in biannual models that estimated differences between participating and non-participating regions for each six-month time period in our dataset. In these models, the six months prior to the start of the trial (from April to September 2021, months −6 to −1) served as the reference period.

Bonferroni correction for multiple comparisons was used to assess significance of analytic results for each secondary group of cancer types. Significance is not assessed, and p-values are not reported, for exploratory analyses of individual cancer types; point estimates and 95% confidence intervals are provided instead. Analyses were performed in R version 4.5.2 (R Foundation for Statistical Computing).

We used eight alternative model specifications to examine the robustness of our findings including analyses that: excluded September to November 2021 as a phase-in period; used the number of hospital beds occupied by patients with confirmed COVID-19 per 100,000 population as an alternate covariate to control for time- and region-varying differences in pandemic-related strain on the healthcare system; included no covariate; applied propensity score weights to account for regional imbalances in population size and number of healthcare staff; estimated an unweighted regression; used wild bootstrapping to generate standard errors to account for limited clusters (regions); used the alternative 14-, 42-, and 62-day cutoffs to calculate diagnostic delay rates; and used average waiting times calculated by mid-point imputation as an alternative outcome specification (appendix methods 2).

Our quasi-experimental difference-in-differences event study design relies on the assumption that, in the absence of exposure to the NHS-Galleri trial, trends in outcomes would be similar for participating and non-participating regions. We visually assess potential deviations from this assumption by examining diagnostic delay rates in the six months preceding the start of the trial (fig 1). Unadjusted rates of diagnostic delays and their trajectories over time were similar across participating and non-participating regions for high-detection cancer types during the pre-trial period. Delay rates in participating regions were slightly higher for the group of low-detection cancer types.

**Figure 1.**
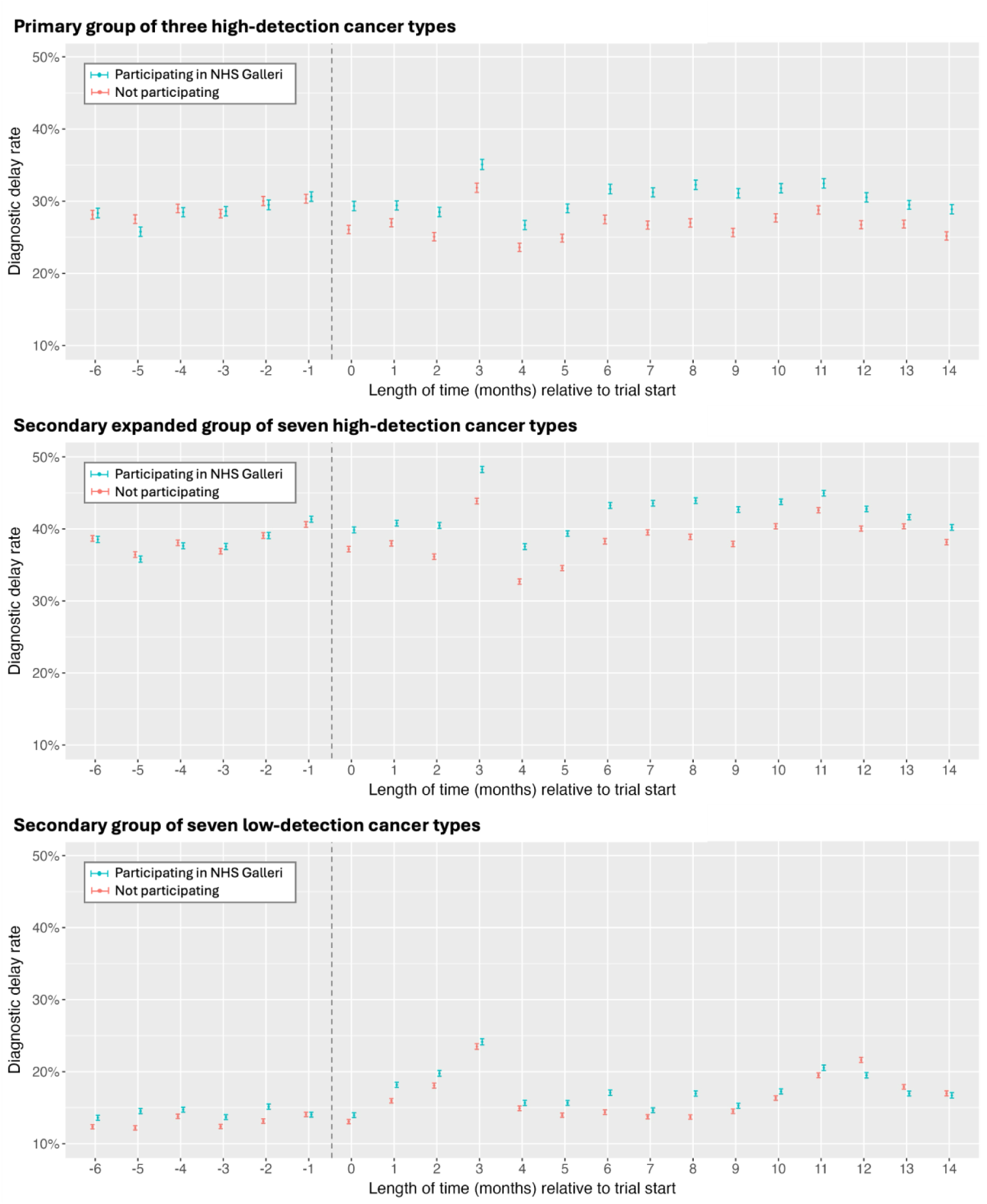
Unadjusted diagnostic delay rates for regions participating in the NHS-Galleri trial compared to non-participating regions during the first half of the study period. The vertical dashed line represents the start of the trial, with the area to the left depicting the six months (months −6 to −1) of the pre-trial baseline period. Months 0 to 14 depict the initial period of participating regions’ exposure to the trial. Diagnostic delay rates are shown as point estimates and 95% confidence intervals.

### Patient and public involvement

No patients or members of the public were involved in developing, conducting, or writing up the results of this study. We did not use individual-level data in our study, which focused instead on data on provider-level referral counts that NHS England has made publicly available for download.

## Results

### Study Sample and Baseline Characteristics

The NHS England Provider-based Cancer Waiting Times dataset reported 10,321,370 patient referrals for suspected cancer during the study period. We excluded 470,084 of these referrals, including 450,392 referrals for non-cancer symptoms or missing information on suspected tumor site and 19,692 referrals from providers serving multiple regions (appendix table 2). Following data cleaning, our study sample included 9,646,631 cancer referrals with suspected tumor site identified (appendix figure 1). 1,875,236 of these referrals were for the primary high-detection group of cancer types, 5,532,510 were for the secondary expanded high-detection group, and 4,114,121 were for the secondary low-detection group of seven cancer types.

The eight regions participating in the NHS-Galleri trial, considered as a group, contained fewer health organization providers and staff than the thirteen non-participating regions (table 1). This corresponds to the smaller patient population served by participating regions as compared to non-participating regions. Population age structure, sex ratio, cancer prevalence, and diagnostic referral rates for cancer were very similar across the two groups.

### Diagnostic Delay Rates

In the first six months after the start of the NHS-Galleri trial, the unadjusted diagnostic delay rate for the primary high-detection group in regions participating in the trial increased from 28.6% to 29.6%, compared to a decrease in non-participating regions from 28.9% to 26.3% (table 3; adjusted difference-in-differences 3.42 percentage points, 95% CI 1.89-4.95, p<0.001, relative increase of 12% from the pre-trial baseline 28.6). Delay rates for the primary group of cancer types remained differentially higher over the next six-month period (adjusted difference-in-differences 4.75 percentage points, 95% CI 1.85-7.66, p=0.003, relative increase of 17% from pre-trial baseline 28.6). Adjusted difference-in-differences correspond to an estimated additional 9,591 referrals experiencing diagnostic delay in the first year of the trial (appendix table 3). Differences receded in the second and third years of the trial (table 3 and fig 2).

**Figure 2.**
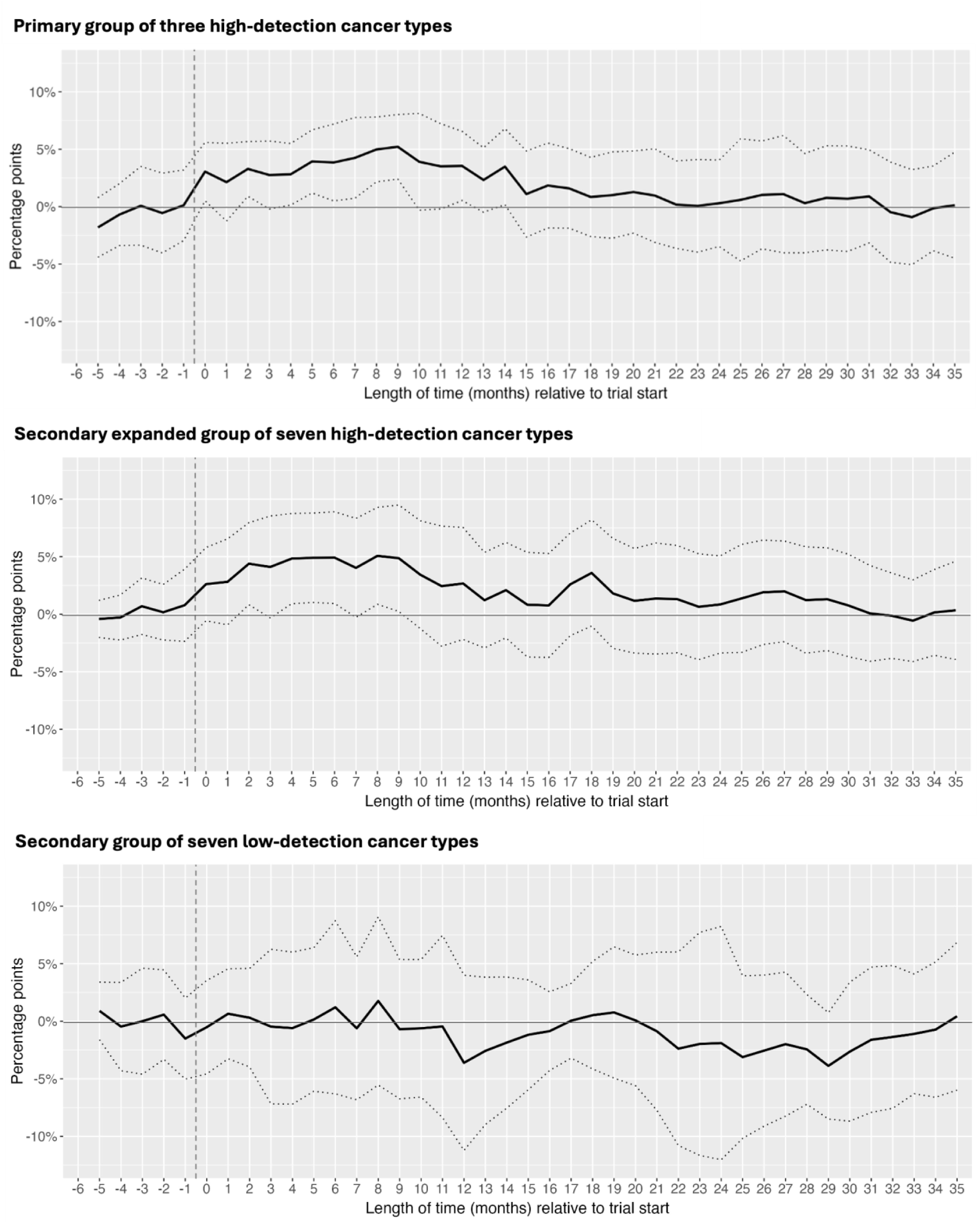
Adjusted differential change in diagnostic delay rates for regions participating in the trial compared to non-participating regions, by month. The vertical dashed line represents the start of the NHS-Galleri trial, with month 0 representing the first month (October 2021) following trial start. The dotted outer lines represent the upper and lower bounds of the 95% CI for each monthly estimate. Monthly estimates are calculated relative to the first month available in our dataset (April 2021, month −6) which served as the reference period. The model includes month and region fixed effects as well as the average percentage of healthcare staff absent in each region in each month. Regressions are weighted by referral volume and include clustered standard errors at the regional level.

We found a significant increase in delay rates for the secondary expanded group of high-detection cancer types in participating regions during the first six months of the trial (table 2; adjusted difference-in-differences 3.73 percentage points, 95% CI 0.83-6.62, p=0.014, significant at Bonferroni-adjusted threshold of p < 0.025). Differences were not significant in subsequent six-month periods. No significant differences in delay rates were observed for the secondary group of low-detection cancer types in the first six-months after trial start (adjusted difference-in-differences 0.03 percentage points, 95% CI −5.07-5.13, p=0.990) or subsequent periods.

**Table 1.** Characteristics of participating and non-participating regions prior to the start of the NHS-Galleri trial.

|  | Participating (n=8) | Not participating (n=13) |
| --- | --- | --- |
| Total no. of providers | 69 | 75 |
| Total no. of staff | 425,957 | 453,353 |
| Total population (1000s) | 25,648 | 30,902 |
| Age, y |  |  |
| <50 | 61.8% | 62.8% |
| 50-64 | 19.3% | 19.0% |
| 65-74 | 10.2% | 9.7% |
| 75-84 | 6.3% | 6.0% |
| 85+ | 2.5% | 2.5% |
| Sex |  |  |
| Female | 50.6% | 50.5% |
| Male | 49.4% | 49.5% |
| Cancer prevalence (per 100,000) | 3,986 | 3,964 |
| Diagnostic referrals for suspected cancer (per 100,000) |  |  |
| All types of suspected cancer | 2,106 | 2,103 |
| Primary high-detection group | 425 | 423 |
| Secondary expanded high-detection group | 1,196 | 1,163 |
| Secondary low-detection group | 901 | 931 |

Note: Number of providers and referral rates are for the six months prior to NHS Galleri start (April – September 2021) from NHS England, Monthly Provider Based Data and Summaries.<sup>28</sup> Six providers included in the data covering the full study period were not recorded as referring any patients during this pre-trial period. Number of staff associated with providers are averaged across the six months prior to the trial and are from NHS Workforce Statistics.<sup>29</sup> Population, age, sex, and cancer prevalence statistics reflect data from December 31, 2020 and are from NHS England, Cancer Prevalence Statistics England 2020.<sup>39</sup>

**Table 2.** Adjusted differential change in diagnostic delay rates for regions participating in the trial compared to non-participating regions.

| Adjusted difference-in-differences, percentage points (95% CI) |  |  |  |  |  |  |
| --- | --- | --- | --- | --- | --- | --- |
| Suspected Cancer Type | Months 0 to 5 | Months 6 to 11 | Months 12 to 17 | Months 18 to 23 | Months 24 to 29 | Months 30 to 35 |
| **Primary high-detection group** | 3.42 (1.89-4.95, p<0.001) | 4.75 (1.85-7.66, p=0.003) | 2.74 (-0.05-5.53, p=0.054) | 1.29 (-2.18-4.76, p=0.447) | 1.20 (-3.24-5.64, p=0.580) | 0.53 (-3.53-4.58, p=0.790) |
| Head & neck | 1.95 (-0.65-4.54) | 3.87 (0.83-6.91) | 2.51 (-0.65-5.66) | 2.05 (-1.48-5.57) | 1.28 (-3.09-5.64) | 0.85 (-2.79-4.48) |
| Lung | 2.16 (-0.00-4.32) | 4.21 (-0.32-8.73) | 0.62 (-4.12-5.36) | 0.03 (-5.23-5.29) | -1.52 (-7.92-4.89) | -3.50 (-9.30-2.30) |
| Upper gastrointestinal | 4.79 (1.07-8.51) | 5.94 (1.58-10.29) | 3.33 (-1.73-8.40) | 1.15 (-5.79-8.10) | 2.25 (-4.28-8.79) | 1.48 (-5.05-8.00) |
| **Secondary expanded high-detection group\*** | 3.73 (0.83-6.62, p=0.014) | 3.91 (0.18-7.65, p=0.041) | 1.54 (-2.35-5.43, p=0.419) | 1.51 (-2.86-5.88, p=0.480) | 1.25 (-3.08-5.58, p=0.553) | -0.06 (-3.56-3.45, p=0.974) |
| Gynaecological cancer | 2.76 (-0.44-5.96) | 1.98 (-3.54-7.50) | 3.87 (-2.22-9.96) | 1.87 (-5.15-8.88) | -0.46 (-5.46-4.54) | -1.26 (-6.81-4.30) |
| Haematological malignancies (excluding acute leukaemia) | 0.19 (-5.80-6.19) | -1.44 (-7.64-4.76) | -3.41 (-11.17-4.35) | -3.17 (-13.64-7.29) | -2.23 (-13.61-9.15) | -5.42 (-15.45-4.61) |
| Lower gastrointestinal | 4.55 (-0.66-9.77) | 3.63 (-2.42-9.69) | -0.23 (-7.41-6.94) | 1.41 (-8.30-11.13) | 2.33 (-6.42-11.08) | 0.21 (-7.01-7.44) |
| Urological malignancies (excluding testicular) | 4.10 (0.39-7.82) | 2.96 (-2.05-7.97) | 0.01 (-5.24-5.26) | 1.63 (-2.56-5.81) | 1.82 (-3.01-6.65) | 0.58 (-4.68-5.84) |
| **Secondary low-detection group** | 0.03 (-5.07-5.13, p=0.990) | 0.20 (-5.90-6.30, p=0.945) | -1.58 (-5.96-2.80, p=0.461) | -0.54 (-6.18-5.09, p=0.843) | -2.51 (-7.73-2.70, p=0.327) | -1.01 (-5.84-3.81, p=0.666) |
| Acute leukaemia | -2.23 (-13.12-8.66) | -8.38 (-28.47-11.70) | 12.29 (-18.36-42.93) | 18.46 (-0.95-37.88) | 25.74 (7.06-44.42) | 17.08 (-5.07-39.22) |
| Brain/central nervous system tumours | 1.16 (-6.86-9.18) | -0.48 (-7.64-6.67) | -0.29 (-10.75-10.17) | -5.41 (-16.66-5.84) | -6.69 (-16.18-2.80) | 2.00 (-11.00-14.99) |
| Breast | 0.25 (-6.70-7.20) | -1.55 (-9.51-6.41) | -0.96 (-5.75-3.83) | 0.62 (-4.75-5.98) | -1.21 (-7.63-5.20) | -0.79 (-4.99-3.41) |
| Sarcoma | 0.48 (-8.30-9.25) | 4.59 (-3.67-12.85) | 7.97 (-4.13-20.06) | 5.76 (-6.18-17.70) | 12.70 (0.89-24.51) | 11.45 (1.57-21.33) |
| Skin | -1.28 (-7.41-4.84) | 0.35 (-6.16-6.87) | -2.73 (-10.29-4.82) | -2.52 (-10.88-5.84) | -4.40 (-11.63-2.83) | -2.65 (-9.94-4.64) |
| Testicular | 2.38 (-3.91-8.68) | 2.72 (-3.83-9.28) | 0.41 (-6.05-6.87) | 2.38 (-3.96-8.71) | -0.95 (-7.71-5.81) | 1.15 (-5.92-8.21) |
| Other | 9.36 (-8.20-26.92) | 5.07 (-15.55-25.68) | 6.35 (-17.46-30.16) | 8.12 (-11.70-27.94) | 5.30 (-15.70-26.31) | 1.92 (-11.47-15.32) |

Note: Adjusted differential changes are estimated for the six six-month periods following the start of the trial in month 0. Estimates are calculated at the level of six-month periods relative to the period prior to the trial start (April 2021 to September 2021, months -6 to -1) using the difference-in-differences model described in methods. The model includes time period and region fixed effects as well as the average percentage of healthcare staff absent in each region over each time period. Regressions are weighted by referral volume and include clustered standard errors at the regional level.
\*The secondary expanded high-detection group consists of the four listed cancer types plus the three cancer types in the primary group above.

Results from our exploratory analyses of individual cancer types suggest that diagnostic delay rates increased in participating regions for urological malignancies in months 0-5 following trial start, for upper gastrointestinal cancer in months 0-11, for head & neck cancer in months 6-11, for acute leukaemia in months 24-29, and for sarcoma in months 24-35.

### Referral Rates

While referral rates were differentially higher in participating regions for all three groupings of cancer types following the start of the trial, this change was not significant for most groups or time periods (appendix table 4). The only significant increase in referral rates observed was for the secondary expanded high-detection group in the second six-month period of the trial (adjusted difference-in-differences 95 referrals per 100,000, 95% CI 17-173, p=0.019, significant at Bonferroni-adjusted threshold of p < 0.025).

### Additional analyses

Our primary finding of increased diagnostic delays for the primary group of high-detection cancer types in participating regions in the first year of the trial was consistent across all sensitivity analyses and outcome specifications (appendix tables 5-14). The increase in diagnostic delay rates for the primary high-detection group corresponded to longer average wait times from referral to diagnostic resolution in participating regions in the first six-month period (appendix table 14; adjusted difference-in-differences 1.70 days, 95% CI 0.97-2.44, p<0.001) and second six-month period (adjusted difference-in-differences 2.29 days, 95% CI 0.86-3.71, p=0.003) following the start of the trial. Our secondary finding that referral rates were differentially higher in participating regions, though not generally to a significant extent, also was consistent across sensitivity analyses (appendix tables 15-20).

## Discussion

We observed a statistically significant increase in diagnostic delay rates for the primary group of high-detection cancer types in regions participating in the NHS-Galleri trial compared to non-participating regions. These differences persisted through the first year of the trial and then receded in the second and third year of the trial. Referral rates were generally higher following the start of the trial but not to a significant extent.

### Possible Explanations and Implications

Several possible mechanisms could explain these results. First, an increase in referrals for intervention group patients receiving positive Galleri test results may have led to greater strain on diagnostic services in regions participating in the trial. In a previous study, average diagnostic follow-up for each positive Galleri test consisted of 4.0 lab tests, 1.9 imaging tests, and 0.4 procedures (e.g. biopsy, endoscopy).^40^ When health services are utilized near full capacity a small increase in demand can lead to a much larger increase in patients facing delay.^41^ Still, it remains unclear whether an expected 100-400 new referrals due to positive Galleri tests in the first year of the trial (see calculations in appendix methods 3) could plausibly explain the additional 9,591 referrals experiencing delay we estimated during that period.

Second, awareness of the trial due to local media coverage, mass invitation to participate in the trial, or proximity to enrolled patients could have encouraged referral-seeking behaviors broadly across participating regions and thus increased demand for scarce diagnostic services. Third, the trial could have disrupted local health system capacity in other ways, if staff involved in diagnostic service delivery were hired to support the trial or if MCED rollout or data collection added to their workload.

The disappearance of spillover effects by the third year of the trial could be due to regional increases in staff, equipment, or other diagnostic resources made in response to higher delays in the first year. Alternatively, early detection of cancer due to the first round of MCED testing might have reduced referrals in subsequent rounds, or any population-level increase in early-detection seeking behavior at the start of the trial could have attenuated over time.

We do not know how increased delay rates affected individual patients in participating regions, though it is likely that the control group experienced similar delays as other patients receiving usual care. If any group received preferential access to diagnostic services, it was likely intervention group patients, who were triaged and monitored on their diagnostic pathway following positive MCED results by designated trial nurses, referral coordinators, and clinical champions.^36^

Delays in diagnostic resolution can lead to stage migration (diagnosis at more advanced stage) and lower survival rates.^42^ An average increase in diagnostic wait times of around two days is unlikely to affect cancer stage for most patients. The clinical significance of such delays depends on patient presentation: for asymptomatic patients identified through screening, where cancers are detected during a preclinical phase with sojourn times of 1-2 years,^43^ small delays likely have minimal impact; however, for symptomatic patients with faster-progressing cancers such as lung cancer, even short delays can be consequential.^44^ Missed opportunities for diagnosis are also a concern and could have a greater impact on cancer stage at time of diagnosis; in a recent study in England, researchers found that doctors responded to higher delay rates by referring fewer patients.^45^ Delayed or missed diagnosis could negatively affect control group outcomes and lead to overestimation of the Galleri test’s effect on the primary trial objective: a reduction in the incidence of later stage cancer at time of diagnosis.

Healthcare decision-makers need a measure of MCED test effectiveness that accurately reflects the extent to which adopting the test within a health system would benefit the patient population as compared to no adoption. Estimates from the NHS-Galleri trial may not accurately reflect this ideal measure of effectiveness, however. First, if spillover led to a system-wide increase in diagnostic delay rates, this would depress control group outcomes during the trial compared to no adoption. Second, MCED test recipients in the intervention group could also face delayed diagnosis due to spillover during the trial, but to a lesser extent than they would if MCED testing was adopted widely across the health system and led to even greater capacity strain. Both factors would lead to overestimation of MCED benefit during the trial compared to actual benefit if adopted broadly across the NHS. Conversely, if system-wide referral rates increased due to the trial, this could lead to improved control group outcomes and underestimation of MCED benefit.

### Comparison to other studies

Our study presents empirical evidence of a potential spillover effect on region-wide rates of diagnostic delays for less commonly referred cancers during the first year of the NHS-Galleri trial. Our quasi-experimental difference-in-differences event study design is similar to that used by others to analyze the potential for spillover due to constraints on service delivery in trials of social welfare and public health interventions.^13,15^ Our analysis complements existing literature examining capacity constraints, cancer diagnostic delay, and health service disruptions during the COVID-19 pandemic.^11,46,47^

Spillover as a potential source of bias has not been addressed in other articles on the NHS-Galleri trial, which have largely focused on the costs of the Galleri test, its accuracy, and use of cancer stage rather than mortality as the primary trial endpoint.^48,49^ The NHS-Galleri study protocol, which considers other sources of bias that could affect trial results such as contamination or differential attrition, does not address the potential for spillover due to constraints on care delivery.^1^ This reflects a broader omission of this form of spillover in the clinical trials literature, including in guidance on trial design,^16,17^ reporting,^18–20^ ethics,^50,51^ and risk of bias assessment.^21–23^

Recent commentaries have raised the possibility that spillover due to constraints on care delivery could affect clinical trials of diverse interventions—including elective induction of labor, AI-based decision support, and patient navigation for cancer diagnosis—but have not provided new empirical evidence supporting this claim.^12,52^ Our findings begin to address this gap and underscore the importance of considering such spillovers in the design of trials evaluating interventions, such as new MCED screening tests, that affect patient utilization of limited healthcare resources.

### Limitations

Our study has several limitations. First, the cancer alliance regions we analyze were deliberately chosen to participate in the trial which raises the possibility of selection bias and residual confounding. This concern, however, is somewhat assuaged by the similar demographic structure, cancer prevalence, referral rates, and pre-trial delay rates in participating and non-participating regions as well as robustness of our results when controlling for regional differences in COVID-19 health system burden. Second, while our study was motivated by the concern that the Galleri test would increase intervention group utilization of diagnostic services and thus strain diagnostic capacity, the NHS-Galleri trial could also have increased diagnostic delay rates via other mechanisms, such as by diverting resources to directly support the trial or by broadly increasing referral-seeking behavior in the region. Finally, we relied solely on publicly available data, aggregated up to the provider-level, that covered a limited timeframe and contained only a narrow set of information.

Future research could help address these limitations. NHS-Galleri researchers will have access to additional data, including on the timing and volume of referrals prompted by the Galleri test as well as on control group outcomes and access to care. Subgroup analysis could focus on delays faced by patients diagnosed with advanced stage cancer. Microsimulation models developed to support power calculations prior to the trial^53^ could be adapted to examine the extent to which increased diagnostic delays and forgone referrals during the first two years of the trial could have affected intervention and control group outcomes. These analyses could inform robustness checks examining the extent to which spillover-induced diagnostic delays might have affected estimates of Galleri test effectiveness during the trial.

### Conclusion

The clinical utility of an MCED test depends not just on its ability to accurately detect early signs of cancer, but on its effectiveness in improving patient outcomes when integrated into the broader continuum of cancer screening, diagnosis, and treatment. This understanding motivates the NHS-Galleri trial and has led government regulators and research funders in the United States to call for similar pragmatic trials of MCED tests in real-world settings. But even as test effectiveness depends on the healthcare system in which it is delivered, the test’s delivery also might change that system, including by increasing utilization of downstream diagnostic resources and thus reducing their availability. As a result of such spillovers, MCED test effectiveness could differ according to the scale of testing and whether tests are delivered together with a corresponding increase in downstream diagnostic resources.

There is limited awareness of the potential for spillover to affect patient outcomes during pragmatic clinical trials. As researchers begin to analyze the results of the NHS-Galleri trial, they should carefully consider the possibility that spillover has affected their results. Future trials evaluating MCED tests, such as the Vanguard study in the United States,^54^ should be designed from the outset to avoid or detect spillover. The possibility that spillover will negatively affect control group and non-participant outcomes should be assessed by health providers and institutional review boards as they weigh risks associated with a proposed trial. Clinical trials of other interventions that affect utilization of limited healthcare resources should also address the potential for spillover to affect patient outcomes and trial results.

## Contributors

SM conceived of the study and obtained funding. SM and BAG developed the plan for the study. SM, JE, PNL, and BAG performed the data analysis. SM wrote the first draft of the paper. SM, JE, PNL, AU, and BAG contributed to interpretation of results and revision of the manuscript. SM supervised the study and is the guarantor. The corresponding author attests that all listed authors meet authorship criteria and that no others meeting the criteria have been omitted.

## Ethics approval

The study used publicly available data and was considered exempt from review by the institutional review board at RAND.

## Data availability statement

We have published the data and analytic code used in this study in a public, open access repository available at: https://github.com/RANDCorporation/mced-spillover-2025

## Transparency

SM affirms that the manuscript is an honest, accurate, and transparent account of the study being reported; that no important aspects of the study have been omitted; and that any discrepancies from the study as planned have been explained.

