## Supplemental Appendix for "Potential spillover effects on diagnostic delay for cancer during the NHS-Galleri trial: a quasi-experimental difference-in-differences study"

Appendix Table 20. Results of sensitivity analysis that used wild bootstrapping to generate standard errors to account for limited clusters (regions). Adjusted differential change in referral rates for regions participating in the trial compared to non-participating regions. . 45

Appendix Methods 3. Back-of-envelope calculation of expected number of new referrals due to positive Galleri tests in the first year of the trial for the purposes of plausibility checks. ........................................................................................................ 47

Appendix Methods 4. Changes made to analysis plan when compared to preregistration and prior analysis. ....................................................................................... 48

**Appendix Figure 1.** Map of the 21 NHS England cancer alliance regions. The eight regions participating in the NHS-Galleri trial are colored purple.

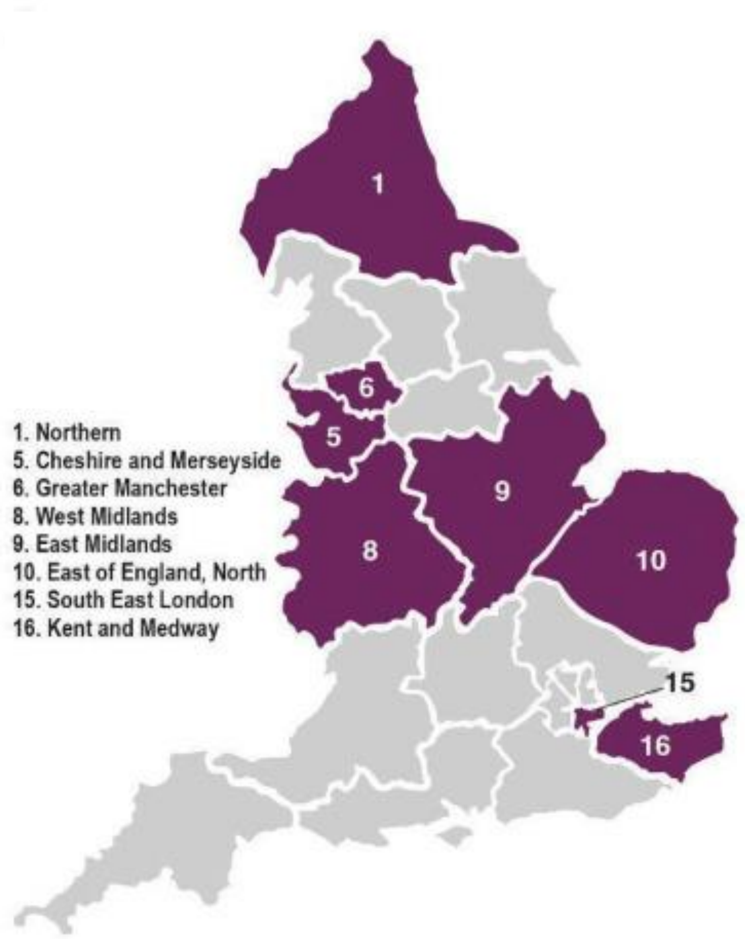

Source: Map image reused under CC-BY license from the NHS-Galleri trial protocol.[1]

**Appendix Table 1.** Results of manual linking of providers to cancer alliance regions. 8 providers listed below from the NHS England Provider-based Cancer Waiting Times dataset provide services across multiple regions.

| Provider Code | Provider Name | Cancer.Alliance | Source/Reasoning |
|---|---|---|---|
| AQK | VERNOVA HEALTHCARE COMMUNITY INTEREST COMPANY | Cheshire and Merseyside Cancer Alliance | Located fully in Cheshire, and thus Cheshire & Merseyside Cancer Alliance (https://www.vernovahealthcare.org/) |
| GEC | RAPID INVESTIGATION SERVICE | Wessex Cancer Alliance | Wessex Cancer Alliance service (https://wessexcanceralliance.nhs.uk/ris-rapid-investigation-service/) |
| NMV | PARTNERSHIPS IN CARE LTD | multiple | "Partnerships in Care is one of the largest and most experienced providers of specialist, secure and step-down care across the UK. With hospitals around the country, 30 years of experience and highly experienced teams, we help patients and commissioners find the right care pathways, without the need for multiple reassessments." https://www.linkedin.com/company/partnerships-in-care/?originalSubdomain=uk |
| NQ7 | MEDWAY COMMUNITY HEALTHCARE | Kent and Medway Cancer Alliance | All locations listed are located in Kent and Medway CA area: https://www.medwaycommunityhealthcare.nhs.uk/contact-us/find-us/ |
| NQT | HCRG CARE LTD | multiple | Independent provider, located across England in multiple cancer alliances. https://www.hcrgcaregroup.com/ |
| NTP | PRACTICE PLUS GROUP | multiple | Independent provider (https://practiceplusgroup.com/) with locations all across England https://practiceplusgroup.com/hospitals/) |
| NV1 | INHEALTH LIMITED | multiple | Independent provider, located across England in multiple cancer alliances) https://www.inhealthgroup.com/locations |
| NVC | RAMSAY HEALTHCARE UK OPERATIONS LIMITED | multiple | Hospitals located across England and Wales. https://www.ramsayhealth.co.uk/hospitals ; "Ramsay Healthcare UK is one of the leading independent healthcare providers in England." https://www.ramsayhealth.co.uk/ |
| NVM | EPSOMEDICAL GROUP | multiple | ("Epsomedical is a small, independent hospital group" "We are not part of any NHS Trust or private hospital group, but have developed as a local provider for Surrey and SW London with units in Epsom and Cobham." https://www.epsomedical.co.uk/language/en-GB/About-Us); Facilities in both Surrey and Sussex Cancer Alliance and SW London / RM Partners CA |
| NYG | SUSSEX COMMUNITY DERMATOLOGY SERVICE | Surrey and Sussex Cancer Alliance | "Sussex Community Dermatology Service has been treating patients in Coastal West Sussex for several years" https://sussexcds.co.uk/services/sussexdermatology/ |
| NYT | ASSURA EAST RIDING LLP | Humber and North Yorkshire Cancer Alliance | independent care provider (Run by HCRG care group, address is "Melton Court, Gibson Lane, Melton, Humberside, HU14 3HH": https://www.cqc.org.uk/location/1-151348139 ; which is "independent provider" https://www.hcrgcaregroup.com/ ; Address located in Humber and North Yorkshire CA https://www.england.nhs.uk/cancer/cancer-alliances-improving-care-locally/) |
| RBZ | NORTHERN DEVON HEALTHCARE NHS TRUST | Somerset, Wiltshire, Avon and Gloucestershire Cancer Alliance | In SWAG Cancer Alliance (https://www.swagcanceralliance.nhs.uk/high-quality-modern-services/) |
| REN | THE CLATTERBRIDGE CANCER CENTRE NHS FOUNDATION TRUST | Cheshire and Merseyside Cancer Alliance | Part of Cheshire and Merseyside Cancer Alliance (https://cmcanceralliance.nhs.uk/news/cancer-centre-one-countrys-top-hospitals) |
| RGD | LEEDS AND YORK PARTNERSHIP NHS FOUNDATION TRUST | multiple | In both Leeds ICS, West Yorkshire ICB, and North Yorkshire ICBs (https://www.leedsandyorkpft.nhs.uk/about-us/our-partnerships/) None are part of MCED Cancer Alliances, but part of two CAs: Humber and North Yorkshire CA and West Yorkshire CA |
| RGM | ROYAL PAPWORTH HOSPITAL NHS | East of England - North Cancer Alliance | Matched to East of England North cancer alliance based on map ([Papworth is north of Cambridge NHS trust which is in North alliance] https://www.canceralliance.co.uk/) |

| | | | |
|---|---|---|---|
| | FOUNDATION TRUST | | |
| RP4 | GREAT ORMOND STREET HOSPITAL FOR CHILDREN NHS FOUNDATION TRUST | North Central London Cancer Alliance | North Central London Cancer Alliance (https://www.nclcanceralliance.nhs.uk/north-central-london-cancer-alliance-map/) |
| RPG | OXLEAS NHS FOUNDATION TRUST | multiple | This provider works in the Kent and Medway CA area and in the South East London CA area "We work out of more than 100 sites across Kent and South East London – here are our main ones:" https://oxleas.nhs.uk/our-sites |
| RPY | THE ROYAL MARSDEN NHS FOUNDATION TRUST | West London Cancer Alliance | ("RM Partners, the west London Cancer Alliance, is leading on the delivery of the recommendations in NHS England's National Cancer Strategy" https://www.royalmarsden.nhs.uk/about-royal-marsden/who-we-are/rm-partners-west-london-cancer-alliance) |
| RRJ | THE ROYAL ORTHOPAEDIC HOSPITAL NHS FOUNDATION TRUST | North Central London Cancer Alliance | (https://www.nclcanceralliance.nhs.uk/north-central-london-cancer-alliance-map/) |
| RT1 | CAMBRIDGESHIRE AND PETERBOROUGH NHS FOUNDATION TRUST | East of England - North Cancer Alliance | CAMBRIDGESHIRE AND PETERBOROUGH NHS FOUNDATION TRUST is part of NHS Cambridgeshire And Peterborough Integrated Care Board which is part of East of England - North CA |
| RT2 | PENNINE CARE NHS FOUNDATION TRUST | Greater Manchester Cancer Alliance | Located in Greater Manchester CA area https://www.penninecare.nhs.uk/ |
| RW6 | PENNINE ACUTE HOSPITALS NHS TRUST | Greater Manchester Cancer Alliance | Combine w other NHS trust data! ("Northern Care Alliance NHS Foundation Trust (NCA) brings together staff and services from Salford Royal NHS Foundation Trust and The Pennine Acute Hospitals NHS Trust. We are now one of the largest NHS providers in the country, working together as a group since 2016 and formally established on 1 October 2021." https://www.northerncarealliance.nhs.uk/about-us) |
| RX2 | SUSSEX PARTNERSHIP NHS FOUNDATION TRUST | Surrey and Sussex Cancer Alliance | Locations are located in the Surrey and Sussex CA area https://www.sussexpartnership.nhs.uk/our-services/hospitals-locations |
| RXY | KENT AND MEDWAY NHS AND SOCIAL CARE PARTNERSHIP TRUST | Kent and Medway Cancer Alliance | Located in the Kent and Medway CA |
| S3H9L | THE HAMPTONS HOSPITAL | East of England - North Cancer Alliance | Located in Peterborough, in the NHS Cambridgeshire and Peterborough ICB and the East of England - North CA area https://thehamptonshospital.com/contact-/#getting-here |

**Appendix Table 2.** Suspected cancer types from NHS England wait times dataset, grouped according to inclusion of high-detection cancers from trial protocol and cancers that are subject to routine screening.

| Suspected Cancer Type | Relationship to high-detection cancers from trial protocol and routine screening |
|---|---|
| **Cancer types that exclusively contain protocol-specified high-detection cancers and are not subject to routine screening:** | |
| Head and neck | Includes one high-detection cancer from trial protocol: Head and neck. Does not include other cancers. |
| Lung | Includes one high-detection cancer from trial protocol: Lung. Does not include other cancers. |
| Upper gastrointestinal | Includes four high-detection cancers from trial protocol: Pancreatic, Liver/bile duct, Stomach, Esophageal. Does not include other cancers. |
| **Cancer types that contain protocol-specified high-detection cancers and either are subject to routine screening or contain additional cancers:** | |
| Gynaecological | Includes one high-detection cancer from trial protocol: Ovarian. Also includes cervical, vulval, and uterine cancers. Cervical cancer is subject to routine NHS screening. |
| Haematological (excluding acute leukaemia) | Includes two high-detection cancers from trial protocol: Myeloma/plasma cell neoplasm, Lymphoma. Also includes chronic leukaemia and other haematological malignancies. |
| Lower gastrointestinal | Includes two high-detection cancers from trial protocol: Colorectal, Anal. Does not include other cancers. Colorectal cancer is subject to routine NHS screening. |
| Urological (excluding testicular) | Includes one high-detection cancer from trial protocol: Bladder. Also includes penile, prostate, and renal cancers. |
| **Cancer types that do not contain any protocol-specified high-detection cancers (Secondary low-detection group):** | |
| Acute leukaemia | Only includes cancers outside of the group of high-detection cancers from trial protocol. |
| Brain/central nervous system | Only includes cancers outside of the group of high-detection cancers from trial protocol. |
| Breast | Only includes cancers outside of the group of high-detection cancers from trial protocol. Breast cancer is subject to routine NHS screening. |
| Sarcoma | Only includes cancers outside of the group of high-detection cancers from trial protocol. |
| Skin | Only includes cancers outside of the group of high-detection cancers from trial protocol. |
| Testicular | Only includes cancers outside of the group of high-detection cancers from trial protocol. |
| Other | Only includes cancers outside of the group of high-detection cancers from trial protocol. |

Note: Suspected cancer types are those provided in the NHS England Provider-based Cancer Waiting Times dataset.[2] Suspected cancer types are mapped to the group of 12 high-detection cancers specified in the trial protocol[1] and to other cancers using the National Cancer Waiting Times Monitoring Dataset Guidance version 12.0.[3]

**Appendix Methods 1.** Simulation modeling to inform study design

For our primary analysis, we analyzed changes in the rate of cancer diagnostic delays in cancer alliance regions that participated in the trial compared to regions that did not participate using a difference-in-differences (DID) framework with an event study design. While recent advances in DID methods have questioned the validity of such models in cases of staggered adoption,[4] the classic DID modeling approach remains valid when all regions are exposed to an intervention during the same time period and in which panel data are balanced.[5]

We performed a simulation analysis to identify the optimal statistical model for use in this analysis. These simulation models were run using the first 2.5 years of data on outcomes and covariates while omitting information on regional treatment assignment (trial participation) status. We compared two-way fixed effects DID models to autoregressive, augmented synthetic controls, and Callaway-Sant'Anna approaches, finding clear evidence that the two-way fixed DID model has robust properties for analyzing this dataset.

In these simulations, using 2.5 years of data (April 2021 – Sep 2023) without linking information on treatment assignment of the regions showed optimal performance of the two-way fixed effects DID model for detecting intervention effects using this dataset. Clustered standard errors at the cancer alliance region level were shown to produce acceptable Type I error rates in our simulations. For the purposes of simulations to determine statistical power, we assumed an effect size of 3 percentage point change in diagnostic delay rates. Simulations showed that we had ~72% power to detect an effect of 3 percentage points change in diagnostic delay rates in this study.

**Appendix Methods 2.** Description of sensitivity analyses.

We used alternative model specifications to examine the robustness of our findings. Each is described below, together with reference to appendix table containing results.

A first sensitivity analysis excluded September, October, and November 2021 as a phase-in period to accommodate uncertainty in timing of exposure to Galleri test rollout across the eight regions participating in the trial. (Appendix Tables 5 & 15)

A second sensitivity analysis dropped the percentage of healthcare staff absent as a covariate in our model. In this analysis, we instead used the number of hospital beds occupied by patients with confirmed COVID-19 per 100,000 population as an alternate covariate to control for time- and region-varying differences in pandemic-related strain on the healthcare system. (Appendix Tables 6 & 16)

A third sensitivity analysis included no covariate in our regression model. (Appendix Tables 7 & 17)

A fourth sensitivity analysis applied propensity score weights to account for regional imbalances in population size and number of healthcare staff. This was conducted following an assessment of the potential utility of using propensity score weights given the constrained number of regions. This assessment relied on data covering the first 2.5 years of our study period (April 2021 – September 2023) on outcomes and covariates, together with additional information on baseline characteristics of each region. We found that the group of participating and non-participating regions were similar on many baseline characteristics (including lagged diagnostic delay rate outcome values, lagged percent of staff absent, population sex ratio and age structure) while there are notable imbalances related to the size of regions in terms of total population size and total number of healthcare staff. (Appendix Tables 8 & 18)

Given the constrained sample size (21 regions) in this analysis, we were unable to produce high quality weights that include more than one or two variables. Thus, we estimated propensity score weights that included population and the number of staff and assessed sensitivity of our models to inclusion of the weights, knowing that they have greatly constrained power but allow us to understand if there might be any lingering bias due to these types of group differences. Balance tables showed the balance was improved for these two covariates after using the estimated propensity score weights. For population and number of staff the unweighted and propensity score weighted effect sizes difference between participating and non-participating regions went from 0.58 to 0.23 and 0.93 to 0.35, respectively.

A fifth sensitivity analysis used unweighted regressions. This stands in contrast to our other analyses of diagnostic delay rates which weighted regressions by the monthly volume of cancer diagnostic referrals. It also stands in contrast to our other analyses of changes in referral rates which weighted regressions by regional population. (Appendix Tables 9 & 19)

A sixth sensitivity analysis used wild bootstrapping to generate robust standard errors that account for the limited number of clusters (21 regions) in our dataset.[6,7] We used Rademacher weights and 999 iterations to generate standard errors using wild cluster bootstrap algorithms. This analysis was performed using the fwildclusterboot R package, version 0.13.0. (Appendix Tables 10 & 20)

A seventh set of sensitivity analyses used alternative specifications of our primary outcome to examine whether estimated changes to diagnostic delay rates are consistent across the alternative 14-, 42-, and 62-day cutoffs provided in our interval-based wait times data. (Appendix Tables 11, 12, 13)

An eighth sensitivity analysis used average waiting times calculated by mid-point imputation as an alternative outcome specification. We calculated this average wait time measure using midpoint imputation for the five intervals provided in our dataset (0–14 days, 15–28 days, 29–42 days, 43–62 days, and more than 62 days), with the final interval treated as 63-82 days for the purposes of imputation. (Appendix Table 14)

**Appendix Figure 2.** Flow diagram depicting number of cancer referrals in original dataset and at each subsequent stage of data cleaning and processing.

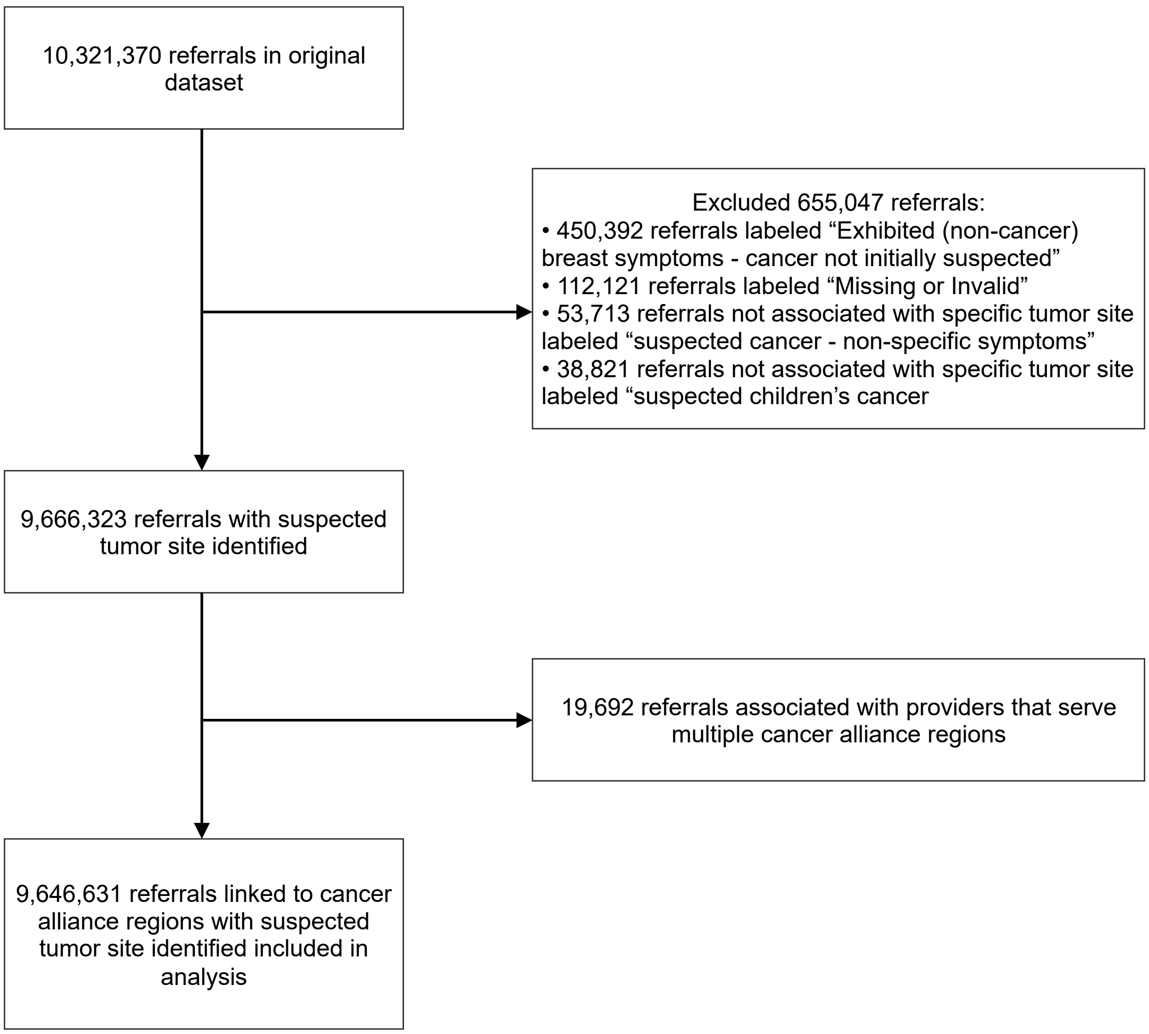

**Appendix Table 3.** Estimated change in number of referrals experiencing diagnostic delay for primary group of high-detection cancers in participating regions.

| | Months 0 to 5 | | | Months 6 to 11 | | |
|---|---|---|---|---|---|---|
| | **Adjusted difference-in-differences in delay rates (percentage points)** | **Total referrals (no.)** | **Change in no. referrals facing delay (no.)** | **Adjusted difference-in-differences in delay rates (percentage points)** | **Total referrals (no.)** | **Change in no. referrals facing delay (no.)** |
| Primary high-detection group | 3.42 | 116,387 | 3,980 | 4.75 | 118,123 | 5,611 |

Note: Estimated change in number of referrals facing diagnostic delays represents the adjusted percentage point difference in delay rates multiplied by the total number of referrals in that time period for participating regions. Only periods with significant differential change in diagnostic delay rates are shown. Table 2 in the main article text shows adjusted differential change in delay rates for all periods.

**Appendix Table 4.** Adjusted differential change in referral rates for regions participating in the trial compared to non-participating regions.

| | Adjusted difference-in-differences, percentage points (95% CI) | | | | | |
|---|---|---|---|---|---|---|
| **Suspected Cancer Type** | **Months 0 to 5** | **Months 6 to 11** | **Months 12 to 17** | **Months 18 to 23** | **Months 24 to 29** | **Months 30 to 35** |
| **Primary high-detection group** | 23.71 (-0.75-48.17, p=0.057) | 12.39 (-6.63-31.40, p=0.189) | 10.32 (-17.36-37.99, p=0.446) | 14.46 (-20.12-49.04, p=0.393) | 19.32 (-21.72-60.36, p=0.338) | 17.43 (-33.57-68.43, p=0.484) |
| Head & neck | 12.44 (-2.03-26.91) | 11.05 (1.40-20.69) | 5.83 (-7.73-19.38) | 11.96 (-2.36-26.29) | 13.75 (-5.89-33.40) | 11.41 (-15.89-38.71) |
| Lung | -0.38 (-5.10-4.35) | -0.94 (-7.72-5.83) | -2.02 (-12.72-8.69) | 0.56 (-14.01-15.12) | 3.27 (-14.97-21.51) | 1.26 (-21.75-24.26) |
| Upper gastrointestinal | 10.17 (-3.23-23.56) | 0.96 (-9.36-11.28) | 6.29 (-7.54-20.12) | 2.66 (-13.75-19.06) | 2.85 (-15.51-21.21) | 3.53 (-16.42-23.48) |
| **Secondary expanded high-detection group*** | 62.53 (-9.64-134.71, p=0.086) | 95.01 (17.21-172.82, p=0.019) | 46.83 (-60.07-153.72, p=0.372) | 79.29 (-37.01-195.59, p=0.170) | 70.58 (-42.72-183.87, p=0.209) | 52.76 (-71.26-176.78, p=0.385) |
| Gynaecological cancer | 7.46 (-8.36-23.27) | 11.02 (-9.35-31.39) | -0.10 (-19.96-19.75) | 11.61 (-14.66-37.88) | 16.11 (-10.39-42.62) | 13.14 (-11.39-37.67) |
| Haematological malignancies (excluding acute leukaemia) | 3.98 (0.54-7.41) | 3.80 (0.31-7.30) | 2.94 (-1.77-7.64) | 4.47 (-0.84-9.77) | 3.65 (-0.90-8.20) | 3.49 (-1.51-8.49) |
| Lower gastrointestinal | 21.81 (-21.04-64.66) | 54.62 (8.08-101.15) | 32.83 (-27.05-92.70) | 27.88 (-26.89-82.65) | 4.01 (-47.94-55.96) | -5.29 (-60.65-50.06) |
| Urological malignancies (excluding testicular) | 7.57 (-4.16-19.30) | 13.35 (-2.82-29.52) | -0.65 (-21.45-20.14) | 19.20 (-3.92-42.33) | 27.62 (-0.29-55.52) | 22.94 (-4.05-49.93) |
| **Secondary low-detection group** | 33.94 (-2.19-70.06, p=0.064) | 33.91 (-36.36-104.17, p=0.326) | 30.97 (-37.27-99.22, p=0.355) | 18.29 (-89.30-125.88, p=0.727) | 8.16 (-103.74-120.05, p=0.881) | -6.45 (-123.28-110.38, p=0.909) |
| Acute leukaemia | 1.03 (0.50-1.56) | -0.05 (-0.19-0.09) | -0.44 (-0.85--0.03) | -0.50 (-1.40-0.39) | 0.22 (-0.42-0.85) | -0.29 (-0.79-0.21) |
| Brain/central nervous system tumours | 1.00 (-1.38-3.38) | 1.43 (-1.54-4.41) | 4.26 (-0.56-9.08) | 0.23 (-2.27-2.73) | 0.28 (-5.42-5.98) | 2.49 (-4.11-9.10) |
| Breast | 8.15 (-20.12-36.43) | 14.83 (-22.12-51.79) | 20.84 (-19.15-60.83) | 12.56 (-28.07-53.19) | 25.58 (-24.46-75.62) | 4.54 (-35.89-44.98) |
| Sarcoma | -2.00 (-8.08-4.08) | 0.08 (-3.88-4.03) | -0.97 (-8.07-6.13) | -11.75 (-28.47-4.97) | -7.29 (-16.76-2.17) | -2.29 (-12.84-8.26) |
| Skin | 30.04 (-5.18-65.25) | 14.99 (-45.20-75.18) | 7.61 (-45.21-60.42) | 7.39 (-83.52-98.30) | -15.40 (-110.97-80.17) | -11.11 (-118.49-96.26) |
| Testicular | 0.42 (-1.38-2.23) | 0.76 (-1.83-3.35) | 0.24 (-1.92-2.41) | 1.43 (-1.38-4.23) | 1.66 (-0.14-3.45) | 1.88 (-0.44-4.21) |
| Other | 5.46 (1.14-9.77) | 10.94 (3.63-18.25) | 12.88 (2.56-23.20) | 6.52 (-0.09-13.12) | 6.26 (-1.81-14.32) | 5.38 (0.08-10.68) |

Note: Adjusted differential changes are estimated for the six six-month periods following the start of the trial in month 0. Estimates are calculated at the level of six-month periods relative to the period prior to the trial start (April 2021 to September 2021, months -6 to -1) using the difference-in-differences model described in methods. The model includes time period and region fixed effects as well as the average percentage of healthcare

staff absent in each region over each time period. Regressions are weighted by population and include clustered standard errors at the regional level.
*The secondary expanded high-detection group consists of the four listed cancer types plus the three cancer types in the primary group above.

**Appendix Table 5.** Results of sensitivity analysis that excluded September to November 2021 as a phase-in period. Adjusted differential change in delay rates for regions participating in the trial compared to non-participating regions.

| | Adjusted difference-in-differences, percentage points (95% CI) | | | | | |
|---|---|---|---|---|---|---|
| **Suspected Cancer Type** | **Months 0 to 5** | **Months 6 to 11** | **Months 12 to 17** | **Months 18 to 23** | **Months 24 to 29** | **Months 30 to 35** |
| **Primary high-detection group** | 3.77 (1.99-5.54, p<0.001) | 4.88 (1.82-7.95, p=0.003) | 2.87 (0.00-5.74, p=0.050) | 1.42 (-2.12-4.97, p=0.412) | 1.33 (-3.20-5.86, p=0.547) | 0.66 (-3.45-4.77, p=0.742) |
| Head & neck | 2.19 (-0.66-5.04) | 3.91 (0.68-7.13) | 2.54 (-0.67-5.75) | 2.06 (-1.67-5.79) | 1.30 (-3.27-5.87) | 0.88 (-2.82-4.58) |
| Lung | 2.41 (-0.27-5.10) | 4.68 (0.09-9.27) | 1.11 (-3.66-5.88) | 0.55 (-4.78-5.89) | -1.00 (-7.26-5.27) | -3.00 (-8.76-2.76) |
| Upper gastrointestinal | 5.15 (0.78-9.52) | 6.07 (1.46-10.68) | 3.48 (-1.81-8.77) | 1.33 (-5.78-8.43) | 2.40 (-4.29-9.10) | 1.62 (-5.04-8.27) |
| **Secondary expanded high-detection group*** | 4.50 (1.04-7.96, p=0.013) | 4.05 (0.12-7.98, p=0.044) | 1.67 (-2.35-5.70, p=0.396) | 1.63 (-2.87-6.13, p=0.458) | 1.38 (-3.10-5.86, p=0.527) | 0.09 (-3.60-3.77, p=0.962) |
| Gynaecological cancer | 2.97 (-1.19-7.12) | 2.47 (-3.06-7.99) | 4.37 (-1.68-10.42) | 2.32 (-4.80-9.44) | 0.01 (-5.14-5.16) | -0.77 (-6.33-4.80) |
| Haematological malignancies (excluding acute leukaemia) | 0.04 (-6.58-6.67) | -2.37 (-8.07-3.33) | -4.33 (-11.68-3.01) | -4.07 (-14.50-6.36) | -3.14 (-14.44-8.17) | -6.34 (-16.22-3.54) |
| Lower gastrointestinal | 5.74 (-0.45-11.93) | 3.57 (-3.07-10.22) | -0.30 (-7.70-7.10) | 1.31 (-8.50-11.12) | 2.26 (-6.62-11.14) | 0.17 (-7.18-7.53) |
| Urological malignancies (excluding testicular) | 5.67 (1.53-9.80) | 3.35 (-1.44-8.15) | 0.41 (-4.63-5.44) | 2.06 (-1.92-6.03) | 2.23 (-2.30-6.75) | 0.97 (-3.98-5.93) |
| **Secondary low-detection group** | -0.36 (-6.82-6.10, p=0.908) | -0.10 (-6.41-6.22, p=0.974) | -1.88 (-6.52-2.76, p=0.408) | -0.84 (-6.79-5.12, p=0.772) | -2.81 (-8.35-2.72, p=0.302) | -1.32 (-6.45-3.82, p=0.598) |
| Acute leukaemia | 1.13 (-13.35-15.62) | -3.45 (-21.97-15.07) | 17.56 (-9.24-44.37) | 24.42 (3.87-44.98) | 32.18 (13.12-51.25) | 22.68 (2.87-42.49) |
| Brain/central nervous system tumours | 1.43 (-7.30-10.16) | -0.56 (-7.32-6.19) | -0.34 (-10.47-9.79) | -5.44 (-16.25-5.38) | -6.54 (-15.47-2.39) | 2.07 (-10.35-14.49) |
| Breast | 0.50 (-8.61-9.60) | -1.64 (-9.95-6.67) | -1.05 (-6.40-4.30) | 0.54 (-5.36-6.44) | -1.30 (-8.26-5.67) | -0.89 (-5.72-3.94) |
| Sarcoma | 0.44 (-9.71-10.59) | 3.10 (-5.33-11.54) | 6.30 (-6.00-18.60) | 5.38 (-6.17-16.92) | 12.09 (0.65-23.52) | 10.66 (0.43-20.90) |
| Skin | -2.72 (-10.14-4.70) | -0.08 (-6.86-6.71) | -3.16 (-11.06-4.74) | -2.95 (-11.80-5.89) | -4.83 (-12.40-2.74) | -3.09 (-10.65-4.47) |
| Testicular | 4.82 (-1.53-11.18) | 3.46 (-2.88-9.81) | 1.17 (-5.05-7.40) | 3.18 (-2.87-9.23) | -0.18 (-6.54-6.18) | 1.92 (-4.85-8.69) |
| Other | 10.99 (-12.30-34.28) | 6.29 (-16.72-29.30) | 6.94 (-20.83-34.71) | 8.37 (-15.26-32.00) | 4.96 (-17.19-27.11) | 1.51 (-15.30-18.32) |

Note: Adjusted differential changes are estimated for the six six-month periods following the start of the trial in month 0. Estimates are calculated at the level of six-month periods (excluding phase-in months) relative to the period prior to the trial start (April 2021 to August 2021, months -6 to -2) using the difference-in-differences model described in methods. The model includes time period and region fixed effects as well as the average

percentage of healthcare staff absent in each region over each time period. Regressions are weighted by referral volume and include clustered standard errors at the regional level.
*The secondary expanded high-detection group consists of the four listed cancer types plus the three cancer types in the primary group above.

**Appendix Table 6.** Results of sensitivity analysis that used the number of hospital beds occupied by patients with confirmed COVID-19 per 100,000 population as an alternate covariate to control for time- and region-varying differences in pandemic-related strain on the healthcare system. Adjusted differential change in delay rates for regions participating in the trial compared to non-participating regions.

| | **Adjusted difference-in-differences, percentage points (95% CI)** | | | | | |
|---|---|---|---|---|---|---|
| **Suspected Cancer Type** | **Months 0 to 5** | **Months 6 to 11** | **Months 12 to 17** | **Months 18 to 23** | **Months 24 to 29** | **Months 30 to 35** |
| **Primary high-detection group** | 3.24 (1.51-4.97, p<0.001) | 4.32 (1.79-6.85, p=0.002) | 2.67 (0.01-5.34, p=0.049) | 1.30 (-1.75-4.34, p=0.385) | 1.27 (-2.73-5.28, p=0.514) | 0.67 (-3.29-4.62, p=0.728) |
| Head & neck | 1.99 (-0.67-4.64) | 3.45 (0.77-6.14) | 2.14 (-0.71-4.99) | 1.27 (-1.85-4.39) | 0.68 (-3.32-4.68) | 0.55 (-3.54-4.64) |
| Lung | 2.16 (-0.00-4.33) | 4.45 (0.10-8.80) | 0.82 (-3.73-5.37) | 0.46 (-4.59-5.50) | -1.14 (-7.50-5.22) | -3.25 (-8.99-2.48) |
| Upper gastrointestinal | 4.25 (-0.20-8.70) | 5.21 (0.91-9.52) | 3.54 (-1.77-8.85) | 1.94 (-5.02-8.89) | 3.00 (-3.52-9.53) | 2.10 (-4.23-8.43) |
| **Secondary expanded high-detection group*** | 3.57 (0.54-6.60, p=0.023) | 3.48 (-0.20-7.17, p=0.063) | 1.43 (-2.39-5.26, p=0.444) | 1.47 (-2.74-5.68, p=0.474) | 1.29 (-2.95-5.54, p=0.532) | 0.06 (-3.58-3.69, p=0.975) |
| Gynaecological cancer | 2.85 (-0.40-6.10) | 1.70 (-3.72-7.12) | 3.69 (-2.40-9.78) | 1.48 (-6.07-9.04) | -0.73 (-6.44-4.98) | -1.39 (-7.42-4.64) |
| Haematological malignancies (excluding acute leukaemia) | 0.37 (-5.58-6.32) | -0.26 (-6.05-5.52) | -2.89 (-10.40-4.63) | -2.35 (-12.53-7.83) | -1.64 (-12.57-9.29) | -5.26 (-15.56-5.04) |
| Lower gastrointestinal | 3.99 (-1.61-9.59) | 2.67 (-3.51-8.85) | -0.23 (-7.23-6.77) | 1.95 (-6.89-10.78) | 2.88 (-5.54-11.30) | 0.81 (-6.29-7.90) |
| Urological malignancies (excluding testicular) | 4.22 (0.94-7.50) | 3.66 (-0.84-8.17) | 0.23 (-5.02-5.48) | 1.88 (-2.56-6.32) | 1.84 (-2.82-6.50) | 0.48 (-4.68-5.64) |
| **Secondary low-detection group** | -0.17 (-5.26-4.93, p=0.946) | -0.06 (-5.73-5.60, p=0.982) | -1.55 (-5.87-2.77, p=0.462) | -0.32 (-5.51-4.88, p=0.900) | -2.30 (-7.29-2.69, p=0.348) | -0.81 (-5.67-4.05, p=0.732) |
| Acute leukaemia | -5.63 (-16.84-5.58) | -13.90 (-32.40-4.60) | 3.66 (-28.91-36.23) | 9.21 (-9.16-27.57) | 21.07 (6.19-35.95) | 12.10 (-5.57-29.76) |
| Brain/central nervous system tumours | 2.62 (-3.35-8.58) | 1.38 (-5.05-7.81) | 3.00 (-6.98-12.99) | -2.13 (-12.22-7.96) | -2.20 (-9.98-5.57) | 6.04 (-5.56-17.65) |
| Breast | 0.72 (-6.49-7.92) | -1.36 (-9.08-6.36) | -1.42 (-5.79-2.96) | -0.32 (-4.87-4.24) | -1.84 (-7.24-3.56) | -1.19 (-4.79-2.42) |
| Sarcoma | 0.31 (-8.79-9.40) | 5.22 (-4.27-14.71) | 8.36 (-4.66-21.39) | 8.37 (-3.20-19.94) | 15.00 (2.53-27.46) | 13.19 (1.41-24.97) |
| Skin | -1.31 (-7.52-4.90) | 0.17 (-6.18-6.52) | -2.23 (-10.24-5.78) | -1.45 (-9.42-6.52) | -3.62 (-10.48-3.23) | -1.92 (-9.14-5.31) |
| Testicular | 2.65 (-3.68-8.99) | 3.37 (-3.95-10.70) | 0.83 (-6.15-7.81) | 2.69 (-4.49-9.87) | -0.77 (-7.78-6.24) | 1.15 (-6.11-8.42) |
| Other | 11.03 (-6.54-28.60) | 8.41 (-10.87-27.68) | 10.30 (-11.46-32.06) | 11.16 (-7.79-30.12) | 7.23 (-14.57-29.02) | 2.88 (-9.40-15.17) |

Note: Adjusted differential changes are estimated for the six six-month periods following the start of the trial in month 0. Estimates are calculated at the level of six-month periods relative to the period prior to the trial start (April 2021 to September 2021, months -6 to -1) using the difference-in-

differences model described in methods. The model includes time period and region fixed effects as well as the number of hospital beds occupied by patients with confirmed COVID-19 per 100,000 population in each region over each time period. Regressions are weighted by referral volume and include clustered standard errors at the regional level.
*The secondary expanded high-detection group consists of the four listed cancer types plus the three cancer types in the primary group above.

**Appendix Table 7.** Results of sensitivity analysis that included no covariate. Adjusted differential change in delay rates for regions participating in the trial compared to non-participating regions.

| | Adjusted difference-in-differences, percentage points (95% CI) | | | | | |
|---|---|---|---|---|---|---|
| **Suspected Cancer Type** | **Months 0 to 5** | **Months 6 to 11** | **Months 12 to 17** | **Months 18 to 23** | **Months 24 to 29** | **Months 30 to 35** |
| **Primary high-detection group** | 3.46 (1.89-5.03, p<0.001) | 4.67 (2.02-7.33, p=0.002) | 2.64 (0.05-5.23, p=0.046) | 1.04 (-1.98-4.07, p=0.481) | 1.00 (-2.98-4.99, p=0.606) | 0.41 (-3.65-4.47, p=0.835) |
| Head & neck | 2.07 (-0.62-4.75) | 3.56 (0.84-6.29) | 2.11 (-0.69-4.91) | 1.15 (-1.73-4.03) | 0.55 (-3.35-4.46) | 0.42 (-3.67-4.51) |
| Lung | 2.12 (0.03-4.21) | 4.38 (-0.15-8.91) | 0.82 (-3.75-5.39) | 0.52 (-4.20-5.23) | -1.07 (-7.13-4.98) | -3.19 (-8.68-2.30) |
| Upper gastrointestinal | 4.74 (0.94-8.54) | 6.02 (1.81-10.24) | 3.44 (-1.68-8.57) | 1.42 (-5.71-8.56) | 2.45 (-4.12-9.02) | 1.59 (-4.96-8.15) |
| **Secondary expanded high-detection group*** | 3.77 (0.88-6.66, p=0.013) | 3.83 (0.06-7.60, p=0.047) | 1.43 (-2.40-5.26, p=0.446) | 1.27 (-3.01-5.54, p=0.543) | 1.07 (-3.09-5.23, p=0.597) | -0.17 (-3.71-3.37, p=0.922) |
| Gynaecological cancer | 2.86 (-0.33-6.05) | 1.84 (-3.66-7.33) | 3.71 (-2.39-9.82) | 1.43 (-6.04-8.90) | -0.79 (-6.36-4.78) | -1.46 (-7.35-4.43) |
| Haematological malignancies (excluding acute leukaemia) | 0.01 (-5.93-5.95) | -1.02 (-6.91-4.87) | -2.87 (-10.28-4.53) | -1.88 (-11.97-8.21) | -1.15 (-12.15-9.85) | -4.78 (-14.83-5.27) |
| Lower gastrointestinal | 4.56 (-0.56-9.67) | 3.63 (-2.57-9.83) | -0.24 (-7.14-6.65) | 1.40 (-7.67-10.46) | 2.32 (-5.83-10.47) | 0.21 (-6.67-7.08) |
| Urological malignancies (excluding testicular) | 3.98 (0.35-7.62) | 3.16 (-1.75-8.07) | 0.23 (-4.92-5.38) | 2.11 (-2.13-6.35) | 2.12 (-2.57-6.80) | 0.73 (-4.54-6.01) |
| **Secondary low-detection group** | 0.02 (-5.10-5.14, p=0.994) | 0.23 (-5.74-6.19, p=0.937) | -1.54 (-5.74-2.65, p=0.451) | -0.46 (-5.43-4.50, p=0.847) | -2.45 (-7.14-2.23, p=0.288) | -0.98 (-5.55-3.58, p=0.658) |
| Acute leukaemia | -5.48 (-16.02-5.06) | -13.88 (-32.26-4.51) | 3.99 (-26.97-34.96) | 9.51 (-6.55-25.57) | 21.00 (6.24-35.75) | 12.36 (-4.24-28.96) |
| Brain/central nervous system tumours | 2.27 (-5.60-10.14) | 0.67 (-5.36-6.70) | 1.11 (-8.53-10.75) | -2.89 (-13.35-7.57) | -3.48 (-11.47-4.50) | 4.91 (-7.56-17.38) |
| Breast | 0.54 (-6.66-7.73) | -1.84 (-9.50-5.81) | -1.37 (-5.86-3.13) | -0.17 (-4.76-4.42) | -1.73 (-7.43-3.97) | -1.11 (-4.89-2.68) |
| Sarcoma | -0.80 (-8.94-7.34) | 4.81 (-3.88-13.50) | 8.18 (-4.17-20.53) | 9.04 (-2.03-20.11) | 15.51 (3.29-27.73) | 13.68 (2.21-25.16) |
| Skin | -1.32 (-7.44-4.80) | 0.63 (-5.87-7.13) | -2.42 (-10.13-5.29) | -1.85 (-9.77-6.08) | -3.95 (-10.59-2.69) | -2.34 (-9.33-4.64) |
| Testicular | 2.29 (-3.96-8.54) | 2.86 (-3.78-9.49) | 0.54 (-6.19-7.27) | 2.78 (-4.47-10.02) | -0.69 (-7.74-6.35) | 1.26 (-6.12-8.64) |
| Other | 11.13 (-6.34-28.60) | 8.59 (-10.35-27.53) | 10.39 (-10.96-31.75) | 11.21 (-7.50-29.93) | 7.27 (-14.42-28.96) | 2.91 (-9.15-14.97) |

Note: Adjusted differential changes are estimated for the six six-month periods following the start of the trial in month 0. Estimates are calculated at the level of six-month periods relative to the period prior to the trial start (April 2021 to September 2021, months -6 to -1) using the difference-in-

differences model described in methods. The model includes time period and region fixed effects. Regressions are weighted by referral volume and include clustered standard errors at the regional level.
*The secondary expanded high-detection group consists of the four listed cancer types plus the three cancer types in the primary group above.

**Appendix Table 8.** Results of sensitivity analysis that applied propensity score weights to account for regional imbalances in population size and number of healthcare staff. Adjusted differential change in delay rates for regions participating in the trial compared to non-participating regions.

| | Adjusted difference-in-differences, percentage points (95% CI) | | | | | |
|---|---|---|---|---|---|---|
| **Suspected Cancer Type** | **Months 0 to 5** | **Months 6 to 11** | **Months 12 to 17** | **Months 18 to 23** | **Months 24 to 29** | **Months 30 to 35** |
| **Primary high-detection group** | 3.93 (2.75-5.12, p<0.001) | 5.84 (3.84-7.84, p<0.001) | 4.83 (2.47-7.19, p<0.001) | 3.44 (0.06-6.83, p=0.047) | 3.29 (-1.66-8.23, p=0.181) | 2.74 (-2.12-7.59, p=0.254) |
| Head & neck | 3.55 (0.26-6.83) | 5.35 (2.27-8.44) | 4.58 (1.02-8.13) | 3.67 (-0.43-7.77) | 3.72 (-0.97-8.40) | 3.27 (-0.99-7.53) |
| Lung | 2.13 (-0.08-4.35) | 5.87 (2.03-9.70) | 0.70 (-3.55-4.94) | 0.68 (-4.85-6.20) | -2.05 (-9.35-5.25) | -3.10 (-10.02-3.83) |
| Upper gastrointestinal | 4.56 (0.89-8.22) | 6.87 (2.15-11.58) | 6.52 (0.93-12.11) | 4.97 (-4.11-14.06) | 5.38 (-3.76-14.53) | 4.85 (-2.67-12.37) |
| **Secondary expanded high-detection group*** | 3.47 (1.80-5.13, p<0.001) | 3.67 (1.45-5.90, p=0.003) | 2.53 (-1.76-6.82, p=0.234) | 2.92 (-2.66-8.50, p=0.288) | 2.62 (-2.86-8.09, p=0.331) | 1.57 (-2.63-5.78, p=0.444) |
| Gynaecological cancer | 1.85 (-0.59-4.30) | 0.97 (-3.27-5.20) | 4.61 (-1.39-10.61) | 0.49 (-7.56-8.54) | 0.64 (-4.60-5.87) | 1.09 (-5.44-7.61) |
| Haematological malignancies (excluding acute leukaemia) | 1.52 (-4.64-7.68) | 1.89 (-4.18-7.96) | 0.10 (-6.76-6.96) | 2.87 (-4.55-10.28) | 3.83 (-4.93-12.59) | -1.07 (-8.77-6.62) |
| Lower gastrointestinal | 3.64 (0.17-7.11) | 2.45 (-1.37-6.28) | 0.75 (-6.78-8.29) | 3.59 (-7.53-14.71) | 2.97 (-7.23-13.18) | 1.35 (-7.48-10.18) |
| Urological malignancies (excluding testicular) | 4.60 (2.18-7.01) | 2.49 (-2.27-7.25) | -1.00 (-6.89-4.89) | 2.23 (-2.92-7.39) | 3.62 (-1.80-9.04) | 2.37 (-3.63-8.37) |
| **Secondary low-detection group** | 2.94 (-1.53-7.41, p=0.186) | 1.82 (-4.08-7.73, p=0.527) | -0.86 (-6.12-4.39, p=0.735) | -0.06 (-5.55-5.42, p=0.981) | -1.21 (-5.84-3.42, p=0.593) | 1.04 (-2.87-4.95, p=0.585) |
| Acute leukaemia | -37.07 (-93.97-19.84) | -22.56 (-81.60-36.48) | 27.07 (-27.05-81.19) | 20.97 (-21.51-63.46) | 24.93 (-41.29-91.15) | 0.19 (-56.99-57.38) |
| Brain/central nervous system tumours | 0.47 (-7.88-8.81) | -3.58 (-13.13-5.96) | 2.01 (-11.09-15.10) | -3.29 (-14.35-7.76) | -5.89 (-12.00-0.21) | 3.08 (-3.66-9.82) |
| Breast | 1.98 (-4.65-8.61) | -1.33 (-9.78-7.13) | 0.09 (-6.42-6.59) | 4.62 (-3.15-12.39) | 3.24 (-4.96-11.43) | 1.64 (-3.08-6.36) |
| Sarcoma | -2.53 (-15.60-10.55) | 12.79 (-8.92-34.49) | 12.25 (-9.96-34.46) | 23.97 (0.50-47.45) | 15.06 (3.16-26.96) | 11.35 (-10.88-33.58) |
| Skin | 3.33 (-1.56-8.22) | 3.72 (-2.23-9.67) | -1.22 (-13.99-11.56) | -4.82 (-18.12-8.49) | -4.99 (-11.80-1.83) | -0.50 (-6.19-5.18) |
| Testicular | -1.57 (-7.51-4.37) | -1.36 (-7.35-4.63) | -3.15 (-10.15-3.86) | 4.43 (-3.56-12.42) | -0.16 (-10.01-9.70) | 1.06 (-9.42-11.53) |
| Other | -10.58 (-42.04-20.88) | 10.17 (-8.08-28.41) | 4.92 (-11.35-21.19) | 8.00 (-15.76-31.75) | 6.36 (-12.14-24.87) | 8.03 (-5.70-21.76) |

Note: Adjusted differential changes are estimated for the six six-month periods following the start of the trial in month 0. Estimates are calculated at the level of six-month periods relative to the period prior to the trial start (April 2021 to September 2021, months -6 to -1) using the difference-in-differences model described in methods. The model includes time period and region fixed effects as well as the average percentage of healthcare staff absent in each region over each time period. Regressions are weighted by propensity score weights and include clustered standard errors at the regional level.
*The secondary expanded high-detection group consists of the four listed cancer types plus the three cancer types in the primary group above.

**Appendix Table 9.** Results of sensitivity analysis that estimated an unweighted regression. Adjusted differential change in delay rates for regions participating in the trial compared to non-participating regions.

| | Adjusted difference-in-differences, percentage points (95% CI) | | | | | |
|---|---|---|---|---|---|---|
| **Suspected Cancer Type** | **Months 0 to 5** | **Months 6 to 11** | **Months 12 to 17** | **Months 18 to 23** | **Months 24 to 29** | **Months 30 to 35** |
| **Primary high-detection group** | 3.64 (1.58-5.70, p=0.001) | 5.73 (2.50-8.95, p=0.001) | 3.59 (0.36-6.82, p=0.031) | 2.19 (-1.58-5.96, p=0.240) | 2.96 (-2.15-8.07, p=0.241) | 1.82 (-2.43-6.06, p=0.383) |
| Head & neck | 2.17 (-0.34-4.67) | 4.68 (1.37-7.98) | 3.09 (-0.45-6.63) | 2.22 (-1.56-6.00) | 2.52 (-2.12-7.17) | 1.75 (-2.18-5.68) |
| Lung | 2.18 (0.12-4.23) | 4.43 (0.64-8.22) | 1.33 (-3.22-5.88) | 1.15 (-3.52-5.82) | -0.44 (-6.57-5.69) | -2.17 (-7.69-3.34) |
| Upper gastrointestinal | 5.54 (1.64-9.45) | 7.94 (2.62-13.25) | 4.93 (-0.24-10.09) | 3.23 (-4.25-10.71) | 5.37 (-2.86-13.60) | 3.92 (-2.96-10.81) |
| **Secondary expanded high-detection group*** | 3.80 (0.18-7.42, p=0.041) | 3.87 (-1.02-8.75, p=0.114) | 1.53 (-3.17-6.23, p=0.504) | 1.53 (-3.48-6.54, p=0.532) | 2.23 (-2.48-6.94, p=0.334) | 0.21 (-3.26-3.68, p=0.901) |
| Gynaecological cancer | 3.01 (-0.83-6.85) | 0.81 (-5.17-6.79) | 2.32 (-3.70-8.34) | -0.12 (-7.69-7.45) | -1.72 (-7.07-3.63) | -3.14 (-8.94-2.65) |
| Haematological malignancies (excluding acute leukaemia) | -2.13 (-8.53-4.26) | -2.90 (-8.07-2.26) | -3.81 (-10.03-2.41) | -1.79 (-8.96-5.38) | -0.35 (-8.79-8.08) | -3.66 (-11.39-4.07) |
| Lower gastrointestinal | 4.87 (-1.32-11.05) | 3.76 (-4.08-11.60) | 0.33 (-8.31-8.96) | 1.83 (-8.69-12.36) | 4.39 (-4.33-13.12) | 1.17 (-5.95-8.29) |
| Urological malignancies (excluding testicular) | 3.98 (-0.81-8.77) | 2.63 (-3.74-9.01) | -0.84 (-6.83-5.15) | 0.59 (-3.74-4.93) | 1.70 (-4.01-7.41) | 0.44 (-5.68-6.57) |
| **Secondary low-detection group** | 0.74 (-4.39-5.86, p=0.767) | 1.48 (-4.77-7.73, p=0.627) | -0.91 (-5.59-3.77, p=0.688) | 0.55 (-3.95-5.06, p=0.801) | -1.32 (-6.01-3.38, p=0.565) | 0.17 (-3.78-4.12, p=0.930) |
| Acute leukaemia | -18.99 (-72.52-34.55) | -29.34 (-87.78-29.09) | 20.62 (-37.24-78.49) | 11.04 (-35.86-57.93) | 11.78 (-62.15-85.71) | -5.97 (-68.26-56.31) |
| Brain/central nervous system tumours | 1.16 (-7.45-9.76) | -3.06 (-10.24-4.11) | -0.27 (-9.84-9.29) | -4.56 (-15.44-6.31) | -8.85 (-17.00--0.69) | 2.01 (-7.77-11.79) |
| Breast | 1.15 (-6.02-8.32) | 0.52 (-7.84-8.89) | 0.26 (-4.01-4.53) | 1.21 (-3.46-5.88) | 0.04 (-6.48-6.57) | -0.32 (-4.16-3.52) |
| Sarcoma | -1.70 (-10.27-6.87) | 12.21 (-6.51-30.92) | 9.23 (-5.75-24.20) | 21.02 (-2.71-44.74) | 9.87 (-3.14-22.87) | 1.03 (-21.32-23.38) |
| Skin | -1.00 (-6.68-4.69) | 1.05 (-5.39-7.49) | -1.86 (-10.43-6.71) | -0.50 (-6.94-5.94) | -2.62 (-8.20-2.97) | -0.70 (-6.72-5.31) |
| Testicular | 0.92 (-4.47-6.32) | 1.73 (-4.89-8.35) | -1.80 (-8.90-5.30) | 2.05 (-6.27-10.37) | -1.27 (-10.05-7.50) | 0.29 (-7.25-7.83) |
| Other | -7.87 (-32.02-16.28) | 5.38 (-17.11-27.88) | 4.31 (-18.95-27.57) | 7.25 (-19.40-33.89) | 10.12 (-9.47-29.72) | -0.28 (-17.10-16.54) |

Note: Adjusted differential changes are estimated for the six six-month periods following the start of the trial in month 0. Estimates are calculated at the level of six-month periods relative to the period prior to the trial start (April 2021 to September 2021, months -6 to -1) using the difference-in-

differences model described in methods. The model includes time period and region fixed effects as well as the average percentage of healthcare staff absent in each region over each time period. Regressions are unweighted and include clustered standard errors at the regional level.
*The secondary expanded high-detection group consists of the four listed cancer types plus the three cancer types in the primary group above.

**Appendix Table 10.** Results of sensitivity analysis that used wild bootstrapping to generate standard errors to account for limited clusters (regions). Adjusted differential change in delay rates for regions participating in the trial compared to non-participating regions.

| | Adjusted difference-in-differences, percentage points (95% CI) | | | | | |
|---|---|---|---|---|---|---|
| **Suspected Cancer Type** | **Months 0 to 5** | **Months 6 to 11** | **Months 12 to 17** | **Months 18 to 23** | **Months 24 to 29** | **Months 30 to 35** |
| **Primary high-detection group** | 3.42 (2.01-4.82, p<0.001) | 4.75 (2.13-7.83, p<0.001) | 2.74 (0.19-5.58, p=0.041) | 1.29 (-2.07-5.38, p=0.443) | 1.20 (-2.94-6.18, p=0.560) | 0.53 (-3.32-4.54, p=0.827) |
| Head & neck | 1.95 (-0.95-4.36) | 3.87 (1.13-7.12) | 2.51 (-0.66-5.84) | 2.05 (-1.18-6.19) | 1.28 (-3.27-6.07) | 0.85 (-2.80-4.92) |
| Lung | 2.16 (-0.21-4.51) | 4.21 (-0.38-8.96) | 0.62 (-4.47-5.93) | 0.03 (-5.81-5.42) | -1.52 (-9.12-4.94) | -3.50 (-9.43-2.65) |
| Upper gastrointestinal | 4.79 (1.08-8.55) | 5.94 (2.03-10.42) | 3.33 (-1.21-8.50) | 1.15 (-5.02-8.13) | 2.25 (-4.15-9.52) | 1.48 (-5.24-8.51) |
| **Secondary expanded high-detection group*** | 3.73 (1.13-6.49, p=0.010) | 3.91 (0.50-7.45, p=0.026) | 1.54 (-2.50-5.36, p=0.378) | 1.51 (-2.77-6.06, p=0.485) | 1.25 (-2.89-5.87, p=0.538) | -0.06 (-3.55-3.57, p=0.985) |
| Gynaecological cancer | 2.76 (-0.30-5.70) | 1.98 (-3.62-7.37) | 3.87 (-2.89-10.02) | 1.87 (-6.35-8.74) | -0.46 (-5.72-4.38) | -1.26 (-7.43-3.81) |
| Haematological malignancies (excluding acute leukaemia) | 0.19 (-6.05-6.44) | -1.44 (-8.20-5.31) | -3.41 (-11.74-4.95) | -3.17 (-14.86-7.04) | -2.23 (-14.79-9.70) | -5.42 (-16.98-3.71) |
| Lower gastrointestinal | 4.55 (-0.45-9.31) | 3.63 (-1.82-9.15) | -0.23 (-7.04-6.60) | 1.41 (-8.26-12.54) | 2.33 (-6.54-11.63) | 0.21 (-6.93-8.03) |
| Urological malignancies (excluding testicular) | 4.10 (0.37-7.51) | 2.96 (-1.71-7.71) | 0.01 (-5.64-4.71) | 1.63 (-2.71-5.78) | 1.82 (-3.30-6.54) | 0.58 (-4.45-5.79) |
| **Secondary low-detection group**** | 0.03 (-5.14-5.23, p=0.988) | 0.20 (-5.65-6.49, p=0.933) | -1.58 (-5.45-2.96, p=0.451) | -0.54 (-6.96-5.36, p=0.867) | -2.51 (-8.60-3.17, p=0.424) | -1.01 (-6.32-3.93, p=0.761) |
| Brain/central nervous system tumours | 1.16 (-7.14-10.79) | -0.48 (-7.64-8.02) | -0.29 (-10.45-13.56) | -5.41 (-18.54-6.45) | -6.69 (-16.35-4.18) | 2.00 (-10.36-18.38) |
| Breast | 0.25 (-6.84-7.10) | -1.55 (-8.59-7.31) | -0.96 (-6.22-4.07) | 0.62 (-4.99-6.18) | -1.21 (-7.43-5.30) | -0.79 (-5.22-3.72) |
| Sarcoma | 0.48 (-14.72-11.32) | 4.59 (-10.38-13.19) | 7.97 (-7.19-24.33) | 5.76 (-7.99-20.46) | 12.70 (0.08-28.20) | 11.45 (0.05-23.14) |
| Skin | -1.28 (-7.34-4.79) | 0.35 (-5.55-7.16) | -2.73 (-9.60-5.41) | -2.52 (-11.03-6.58) | -4.40 (-12.89-3.37) | -2.65 (-10.67-5.05) |
| Testicular | 2.38 (-3.63-9.31) | 2.72 (-4.75-9.30) | 0.41 (-6.74-7.09) | 2.38 (-4.36-8.73) | -0.95 (-8.14-5.38) | 1.15 (-6.11-9.70) |
| Other | 9.36 (-11.18-30.44) | 5.07 (-28.66-29.62) | 6.35 (-25.88-31.52) | 8.12 (-16.71-31.94) | 5.30 (-21.02-36.47) | 1.92 (-18.89-18.62) |

Note: Adjusted differential changes are estimated for the six six-month periods following the start of the trial in month 0. Estimates are calculated at the level of six-month periods relative to the period prior to the trial start (April 2021 to September 2021, months -6 to -1) using the difference-in-differences model described in methods. The model includes time period and region fixed effects as well as the average percentage of healthcare

staff absent in each region over each time period. Regressions are weighted by referral volume and use wild bootstrapping to generate standard errors to account for limited clusters (regions).
*The secondary expanded high-detection group consists of the four listed cancer types plus the three cancer types in the primary group above.
**We were unable to produce results of wild bootstrap analysis for individual analysis of acute leukaemia, which had the smallest number of referrals of all individual cancer types.

**Appendix Table 11.** Results of sensitivity analysis that used the alternative 14-day cutoff to calculate diagnostic delay rates. Adjusted differential change in delay rates for regions participating in the trial compared to non-participating regions.

| | Adjusted difference-in-differences, percentage points (95% CI) | | | | | |
|---|---|---|---|---|---|---|
| **Suspected Cancer Type** | **Months 0 to 5** | **Months 6 to 11** | **Months 12 to 17** | **Months 18 to 23** | **Months 24 to 29** | **Months 30 to 35** |
| **Primary high-detection group** | 3.85 (1.87-5.83, p<0.001) | 4.00 (0.52-7.48, p=0.026) | 2.40 (-1.38-6.18, p=0.201) | 0.33 (-4.20-4.87, p=0.879) | -0.28 (-6.35-5.80, p=0.925) | -1.24 (-5.85-3.36, p=0.579) |
| Head & neck | 2.91 (0.37-5.46) | 5.35 (1.99-8.72) | 3.70 (-0.08-7.48) | 2.66 (-2.29-7.62) | 1.40 (-3.08-5.89) | -0.38 (-4.32-3.56) |
| Lung | 1.10 (-4.51-6.70) | 3.40 (-2.52-9.33) | 1.33 (-6.91-9.58) | -0.24 (-9.30-8.82) | -3.29 (-11.96-5.37) | -5.54 (-12.66-1.58) |
| Upper gastrointestinal | 5.13 (1.63-8.63) | 3.00 (-1.41-7.41) | 0.89 (-5.29-7.07) | -1.78 (-9.73-6.17) | -0.82 (-10.30-8.65) | -0.62 (-8.87-7.63) |
| **Secondary expanded high-detection group*** | 2.58 (0.67-4.50, p=0.011) | 2.82 (0.00-5.64, p=0.050) | 1.13 (-1.74-4.01, p=0.420) | 0.01 (-3.25-3.27, p=0.996) | -0.32 (-3.84-3.21, p=0.853) | -0.83 (-3.84-2.19, p=0.574) |
| Gynaecological cancer | 1.10 (-1.02-3.22) | 0.85 (-3.67-5.37) | 2.97 (-1.42-7.37) | 0.17 (-5.34-5.68) | -1.27 (-6.29-3.75) | -1.23 (-7.18-4.73) |
| Haematological malignancies (excluding acute leukaemia) | 0.40 (-2.42-3.22) | -1.25 (-5.93-3.43) | -3.44 (-8.51-1.64) | -4.35 (-12.86-4.16) | -1.49 (-9.72-6.73) | -4.51 (-12.19-3.16) |
| Lower gastrointestinal | 2.38 (-0.40-5.16) | 2.30 (-0.74-5.33) | 0.61 (-3.54-4.75) | 0.05 (-6.22-6.32) | 1.51 (-4.87-7.89) | 1.00 (-4.45-6.44) |
| Urological malignancies (excluding testicular) | 2.50 (-0.50-5.50) | 1.44 (-2.63-5.52) | -2.98 (-7.42-1.46) | -1.57 (-5.84-2.69) | -2.60 (-7.04-1.85) | -2.07 (-6.92-2.78) |
| **Secondary low-detection group** | 1.30 (-6.21-8.82, p=0.721) | -0.62 (-9.27-8.03, p=0.882) | 0.48 (-7.18-8.14, p=0.897) | -3.44 (-12.31-5.42, p=0.427) | -3.85 (-12.38-4.68, p=0.358) | -4.91 (-13.31-3.49, p=0.237) |
| Acute leukaemia | -1.88 (-18.84-15.07) | -16.21 (-46.23-13.81) | -0.87 (-32.57-30.83) | 15.32 (-6.69-37.33) | 7.86 (-11.35-27.07) | -1.35 (-28.15-25.45) |
| Brain/central nervous system tumours | -0.37 (-7.34-6.59) | -4.76 (-19.92-10.39) | 3.59 (-10.90-18.08) | -8.35 (-28.24-11.55) | -11.76 (-26.67-3.16) | -4.52 (-25.69-16.65) |
| Breast | 3.77 (-8.65-16.18) | 0.31 (-14.73-15.36) | 5.64 (-7.67-18.96) | 2.48 (-9.30-14.26) | 0.64 (-12.79-14.07) | -1.61 (-11.60-8.39) |
| Sarcoma | -1.44 (-10.45-7.57) | 4.85 (-6.25-15.95) | 6.63 (-8.43-21.69) | 3.85 (-10.70-18.40) | 11.49 (-1.37-24.36) | 13.24 (0.27-26.20) |
| Skin | -2.42 (-9.97-5.13) | -2.76 (-10.69-5.18) | -4.73 (-13.95-4.48) | -9.22 (-20.12-1.68) | -8.96 (-19.95-2.03) | -9.29 (-21.28-2.70) |
| Testicular | 1.85 (-6.44-10.13) | -0.15 (-10.20-9.91) | 1.01 (-9.59-11.61) | 4.60 (-2.85-12.04) | 1.71 (-7.97-11.39) | 3.73 (-7.46-14.93) |
| Other | 4.83 (-5.40-15.06) | 13.60 (2.51-24.68) | 10.89 (-7.43-29.22) | 7.08 (-10.75-24.91) | 7.50 (-8.21-23.21) | 15.34 (3.32-27.37) |

Note: Adjusted differential changes are estimated for the six six-month periods following the start of the trial in month 0. Estimates are calculated at the level of six-month periods relative to the period prior to the trial start (April 2021 to September 2021, months -6 to -1) using the difference-in-differences model described in methods. The model includes time period and region fixed effects as well as the average percentage of healthcare

staff absent in each region over each time period. Regressions are weighted by referral volume and include clustered standard errors at the regional level.
*The secondary expanded high-detection group consists of the four listed cancer types plus the three cancer types in the primary group above.

**Appendix Table 12.** Results of sensitivity analysis that used the alternative 42-day cutoff to calculate diagnostic delay rates. Adjusted differential change in delay rates for regions participating in the trial compared to non-participating regions.

| | Adjusted difference-in-differences, percentage points (95% CI) | | | | | |
|---|---|---|---|---|---|---|
| **Suspected Cancer Type** | **Months 0 to 5** | **Months 6 to 11** | **Months 12 to 17** | **Months 18 to 23** | **Months 24 to 29** | **Months 30 to 35** |
| **Primary high-detection group** | 2.59 (1.31-3.87, p<0.001) | 3.77 (1.53-6.00, p=0.002) | 2.38 (0.05-4.72, p=0.046) | 1.79 (-1.01-4.60, p=0.197) | 1.45 (-1.91-4.81, p=0.378) | 1.09 (-2.06-4.25, p=0.478) |
| Head & neck | 2.15 (-0.24-4.53) | 2.90 (0.16-5.64) | 1.99 (-0.70-4.68) | 2.21 (-0.48-4.90) | 1.39 (-2.03-4.81) | 1.17 (-1.71-4.05) |
| Lung | 0.74 (-0.63-2.12) | 2.92 (0.75-5.09) | 1.71 (-1.20-4.62) | 1.63 (-1.64-4.90) | 0.59 (-3.03-4.22) | -0.65 (-4.38-3.08) |
| Upper gastrointestinal | 3.23 (0.48-5.99) | 5.07 (1.39-8.75) | 2.97 (-0.93-6.88) | 1.82 (-3.69-7.34) | 2.15 (-2.92-7.22) | 1.65 (-3.38-6.69) |
| **Secondary expanded high-detection group*** | 3.05 (0.55-5.54, p=0.019) | 3.84 (0.51-7.18, p=0.026) | 1.67 (-1.67-5.01, p=0.309) | 1.78 (-1.80-5.36, p=0.311) | 1.32 (-2.35-4.99, p=0.461) | 0.38 (-2.79-3.54, p=0.807) |
| Gynaecological cancer | 1.86 (-0.84-4.57) | 1.42 (-3.18-6.02) | 1.77 (-2.98-6.52) | -0.06 (-5.34-5.21) | -0.88 (-4.38-2.62) | -1.09 (-4.60-2.42) |
| Haematological malignancies (excluding acute leukaemia) | -0.07 (-4.32-4.18) | 0.19 (-4.93-5.31) | -2.96 (-8.44-2.52) | -1.11 (-8.79-6.58) | -1.20 (-11.31-8.92) | -4.42 (-12.15-3.31) |
| Lower gastrointestinal | 3.95 (-0.49-8.40) | 5.07 (-0.09-10.23) | 1.06 (-5.15-7.27) | 2.86 (-5.36-11.08) | 1.80 (-5.60-9.19) | 0.20 (-6.17-6.58) |
| Urological malignancies (excluding testicular) | 3.44 (0.43-6.45) | 2.52 (-2.08-7.12) | 1.09 (-3.66-5.83) | 1.61 (-1.73-4.94) | 2.88 (-0.94-6.71) | 1.54 (-2.49-5.57) |
| **Secondary low-detection group** | -0.85 (-3.46-1.76, p=0.504) | -0.96 (-4.03-2.12, p=0.524) | -0.94 (-4.07-2.20, p=0.541) | 0.14 (-2.97-3.24, p=0.928) | -0.66 (-3.78-2.46, p=0.666) | -1.17 (-3.96-1.61, p=0.391) |
| Acute leukaemia | -2.98 (-12.26-6.29) | -9.47 (-17.03--1.91) | 7.04 (-8.80-22.88) | 12.56 (-3.56-28.68) | 18.72 (-0.49-37.93) | 16.82 (6.25-27.39) |
| Brain/central nervous system tumours | 2.90 (-2.10-7.89) | 2.40 (-3.95-8.75) | 2.64 (-4.38-9.66) | -1.43 (-8.41-5.56) | -2.16 (-8.73-4.42) | 2.82 (-6.16-11.80) |
| Breast | -1.31 (-3.31-0.68) | -1.33 (-3.45-0.80) | -1.15 (-3.05-0.74) | -0.77 (-2.47-0.93) | -0.49 (-2.35-1.37) | -0.83 (-2.29-0.63) |
| Sarcoma | -1.98 (-8.62-4.65) | 1.31 (-3.59-6.21) | 5.75 (-3.84-15.35) | 2.41 (-4.63-9.44) | 7.96 (0.56-15.35) | 7.03 (1.15-12.91) |
| Skin | -0.77 (-5.37-3.82) | -1.51 (-6.31-3.30) | -1.04 (-7.34-5.26) | 0.18 (-5.18-5.54) | -1.16 (-6.62-4.29) | -2.40 (-7.28-2.48) |
| Testicular | 2.14 (-3.41-7.69) | 2.24 (-2.92-7.39) | 0.90 (-3.13-4.92) | 1.30 (-2.69-5.29) | 0.61 (-3.68-4.90) | -0.23 (-4.09-3.64) |
| Other | 8.06 (-8.51-24.62) | 5.76 (-10.94-22.47) | 3.64 (-15.51-22.79) | 11.72 (-4.38-27.83) | 6.75 (-13.41-26.92) | 1.43 (-8.65-11.52) |

Note: Adjusted differential changes are estimated for the six six-month periods following the start of the trial in month 0. Estimates are calculated at the level of six-month periods relative to the period prior to the trial start (April 2021 to September 2021, months -6 to -1) using the difference-in-differences model described in methods. The model includes time period and region fixed effects as well as the average percentage of healthcare

staff absent in each region over each time period. Regressions are weighted by referral volume and include clustered standard errors at the regional level.
*The secondary expanded high-detection group consists of the four listed cancer types plus the three cancer types in the primary group above.

**Appendix Table 13.** Results of sensitivity analysis that used the alternative 62-day cutoff to calculate diagnostic delay rates. Adjusted differential change in delay rates for regions participating in the trial compared to non-participating regions.

| | Adjusted difference-in-differences, percentage points (95% CI) | | | | | |
|---|---|---|---|---|---|---|
| **Suspected Cancer Type** | **Months 0 to 5** | **Months 6 to 11** | **Months 12 to 17** | **Months 18 to 23** | **Months 24 to 29** | **Months 30 to 35** |
| **Primary high-detection group** | 1.13 (0.34-1.92, p=0.007) | 2.01 (0.69-3.34, p=0.005) | 1.12 (-0.23-2.46, p=0.099) | 1.10 (-0.56-2.76, p=0.182) | 0.68 (-1.27-2.63, p=0.477) | 0.20 (-1.72-2.13, p=0.829) |
| Head & neck | 0.59 (-0.65-1.84) | 1.04 (-0.48-2.56) | 0.66 (-1.10-2.42) | 0.90 (-0.49-2.30) | 0.47 (-1.23-2.16) | -0.52 (-2.28-1.24) |
| Lung | 0.61 (-0.25-1.46) | 2.30 (1.29-3.31) | 1.27 (0.05-2.48) | 1.46 (0.08-2.84) | 1.12 (-0.57-2.81) | 0.58 (-1.22-2.39) |
| Upper gastrointestinal | 1.70 (0.03-3.37) | 3.13 (0.49-5.76) | 1.59 (-1.03-4.21) | 1.49 (-2.22-5.19) | 0.96 (-2.53-4.44) | 1.09 (-2.27-4.45) |
| **Secondary expanded high-detection group*** | 1.47 (-0.09-3.03, p=0.064) | 2.43 (0.06-4.80, p=0.045) | 1.23 (-0.93-3.39, p=0.249) | 1.40 (-0.67-3.47, p=0.173) | 0.81 (-1.42-3.04, p=0.457) | 0.21 (-1.81-2.24, p=0.830) |
| Gynaecological cancer | 0.44 (-1.16-2.03) | 0.84 (-1.63-3.31) | 0.12 (-2.57-2.81) | 0.18 (-2.76-3.11) | -0.72 (-2.95-1.51) | -0.64 (-2.49-1.21) |
| Haematological malignancies (excluding acute leukaemia) | 1.03 (-2.06-4.12) | 1.49 (-2.31-5.29) | -1.06 (-4.16-2.04) | -0.82 (-4.46-2.82) | 0.34 (-4.54-5.23) | -0.60 (-4.00-2.80) |
| Lower gastrointestinal | 2.17 (-0.76-5.11) | 3.77 (-0.20-7.74) | 1.81 (-2.40-6.01) | 2.67 (-1.86-7.20) | 1.29 (-3.10-5.68) | 0.55 (-3.16-4.26) |
| Urological malignancies (excluding testicular) | 1.62 (-0.38-3.61) | 1.25 (-2.35-4.85) | 1.08 (-2.61-4.76) | 0.77 (-2.06-3.60) | 1.86 (-0.81-4.54) | 0.75 (-2.62-4.12) |
| **Secondary low-detection group** | -0.06 (-1.37-1.24, p=0.921) | -0.66 (-2.25-0.92, p=0.393) | -0.04 (-1.65-1.56, p=0.958) | 0.54 (-0.84-1.92, p=0.420) | 0.11 (-1.67-1.88, p=0.902) | -0.65 (-2.27-0.96, p=0.409) |
| Acute leukaemia | 5.58 (-1.73-12.89) | 11.98 (7.78-16.17) | 14.65 (6.64-22.65) | 19.94 (11.64-28.24) | 19.39 (7.28-31.50) | 16.83 (11.45-22.21) |
| Brain/central nervous system tumours | 2.10 (-0.82-5.02) | 1.66 (-3.44-6.75) | 2.85 (-2.54-8.24) | 0.90 (-3.72-5.52) | -1.10 (-4.25-2.05) | 0.79 (-4.76-6.34) |
| Breast | -0.36 (-1.01-0.29) | -0.56 (-1.25-0.13) | -0.47 (-1.27-0.33) | -0.34 (-1.01-0.34) | -0.26 (-1.08-0.55) | -0.44 (-1.14-0.27) |
| Sarcoma | -1.51 (-4.48-1.45) | -0.12 (-3.44-3.21) | 1.25 (-5.80-8.30) | -0.35 (-3.43-2.74) | 2.37 (-1.58-6.33) | 3.62 (0.59-6.65) |
| Skin | 0.18 (-2.47-2.82) | -1.06 (-3.77-1.66) | 0.37 (-2.76-3.50) | 1.04 (-1.38-3.45) | 0.42 (-2.80-3.65) | -1.17 (-3.99-1.65) |
| Testicular | 0.84 (-1.45-3.13) | 0.93 (-2.10-3.96) | -0.05 (-2.27-2.17) | -0.42 (-2.45-1.62) | -0.19 (-2.05-1.68) | -0.86 (-2.81-1.10) |
| Other | 3.32 (-2.60-9.24) | 0.30 (-5.85-6.45) | 0.08 (-7.94-8.09) | 4.34 (-3.54-12.22) | 1.36 (-4.75-7.47) | -3.79 (-9.99-2.41) |

Note: Adjusted differential changes are estimated for the six six-month periods following the start of the trial in month 0. Estimates are calculated at the level of six-month periods relative to the period prior to the trial start (April 2021 to September 2021, months -6 to -1) using the difference-in-differences model described in methods. The model includes time period and region fixed effects as well as the average percentage of healthcare

staff absent in each region over each time period. Regressions are weighted by referral volume and include clustered standard errors at the regional level.
*The secondary expanded high-detection group consists of the four listed cancer types plus the three cancer types in the primary group above.

**Appendix Table 14.** Results of sensitivity analysis that used the alternative outcome specification of estimated average wait times calculated using mid-point imputation. Adjusted differential change in wait times (days) from referral to diagnostic resolution for regions participating in the trial compared to non-participating regions.

| | Adjusted difference-in-differences, days (95% CI) | | | | | |
|---|---|---|---|---|---|---|
| **Suspected Cancer Type** | **Months 0 to 5** | **Months 6 to 11** | **Months 12 to 17** | **Months 18 to 23** | **Months 24 to 29** | **Months 30 to 35** |
| **Primary high-detection group** | 1.70 (0.97-2.44, p<0.001) | 2.29 (0.86-3.71, p=0.003) | 1.36 (-0.06-2.78, p=0.060) | 0.75 (-0.96-2.47, p=0.369) | 0.51 (-1.80-2.82, p=0.650) | 0.12 (-1.81-2.05, p=0.899) |
| Head & neck | 1.18 (0.04-2.32) | 2.02 (0.61-3.43) | 1.36 (-0.15-2.86) | 1.23 (-0.14-2.59) | 0.71 (-1.25-2.67) | 0.16 (-1.40-1.72) |
| Lung | 0.71 (-0.55-1.97) | 2.04 (0.41-3.66) | 0.82 (-1.34-2.99) | 0.54 (-1.98-3.06) | -0.36 (-3.21-2.48) | -1.29 (-3.91-1.33) |
| Upper gastrointestinal | 2.30 (0.68-3.93) | 2.75 (0.61-4.90) | 1.42 (-1.04-3.88) | 0.51 (-2.94-3.96) | 0.75 (-2.61-4.12) | 0.62 (-2.55-3.78) |
| **Secondary expanded high-detection group*** | 1.71 (0.36-3.06, p=0.016) | 2.09 (0.23-3.96, p=0.030) | 0.91 (-0.95-2.77, p=0.320) | 0.80 (-1.18-2.77, p=0.410) | 0.52 (-1.52-2.56, p=0.604) | -0.02 (-1.74-1.69, p=0.980) |
| Gynaecological cancer | 0.95 (-0.38-2.28) | 0.81 (-1.72-3.34) | 1.30 (-1.36-3.95) | 0.31 (-2.68-3.30) | -0.54 (-2.69-1.60) | -0.67 (-3.03-1.70) |
| Haematological malignancies (excluding acute leukaemia) | 0.28 (-1.90-2.45) | -0.05 (-2.89-2.78) | -1.69 (-4.76-1.38) | -1.43 (-5.94-3.08) | -0.66 (-5.91-4.58) | -2.28 (-6.57-2.00) |
| Lower gastrointestinal | 2.09 (-0.21-4.39) | 2.46 (-0.16-5.08) | 0.60 (-2.64-3.84) | 1.23 (-3.16-5.61) | 1.11 (-2.92-5.14) | 0.32 (-3.05-3.69) |
| Urological malignancies (excluding testicular) | 1.84 (0.11-3.58) | 1.30 (-1.37-3.97) | -0.03 (-2.84-2.78) | 0.43 (-1.69-2.55) | 0.74 (-1.54-3.02) | 0.19 (-2.30-2.68) |
| **Secondary low-detection group** | 0.04 (-2.19-2.26, p=0.974) | -0.36 (-3.07-2.36, p=0.787) | -0.32 (-2.61-1.97, p=0.775) | -0.44 (-3.12-2.24, p=0.733) | -1.00 (-3.51-1.51, p=0.416) | -1.18 (-3.64-1.28, p=0.327) |
| Acute leukaemia | 0.02 (-5.34-5.39) | -2.74 (-10.09-4.61) | 5.72 (-4.62-16.07) | 10.93 (3.35-18.51) | 11.80 (4.52-19.09) | 8.42 (0.38-16.46) |
| Brain/central nervous system tumours | 1.02 (-1.65-3.69) | -0.02 (-4.29-4.25) | 1.50 (-3.59-6.59) | -2.03 (-7.92-3.86) | -3.23 (-8.02-1.56) | 0.26 (-6.78-7.30) |
| Breast | 0.29 (-2.68-3.25) | -0.51 (-4.17-3.15) | 0.39 (-2.59-3.37) | 0.25 (-2.47-2.97) | -0.21 (-3.28-2.85) | -0.57 (-2.71-1.56) |
| Sarcoma | -0.78 (-4.64-3.07) | 1.55 (-2.23-5.32) | 3.30 (-3.15-9.76) | 1.70 (-3.62-7.03) | 5.27 (0.10-10.45) | 5.44 (0.95-9.93) |
| Skin | -0.63 (-3.64-2.38) | -0.82 (-3.85-2.22) | -1.17 (-4.77-2.43) | -1.45 (-5.15-2.25) | -2.03 (-5.60-1.54) | -2.36 (-6.14-1.42) |
| Testicular | 1.13 (-2.14-4.41) | 0.93 (-2.74-4.59) | 0.35 (-2.97-3.66) | 1.14 (-1.51-3.79) | 0.18 (-3.00-3.36) | 0.49 (-2.76-3.75) |
| Other | 4.04 (-3.08-11.17) | 3.72 (-3.46-10.90) | 3.10 (-6.30-12.51) | 5.02 (-3.31-13.36) | 3.25 (-5.34-11.84) | 1.98 (-2.88-6.84) |

Note: Adjusted differential changes are estimated for the six six-month periods following the start of the trial in month 0. Estimates are calculated at the level of six-month periods relative to the period prior to the trial start (April 2021 to September 2021, months -6 to -1) using the difference-in-

differences model described in methods. The model includes time period and region fixed effects as well as the average percentage of healthcare staff absent in each region over each time period. Regressions are weighted by referral volume and include clustered standard errors at the regional level.
*The secondary expanded high-detection group consists of the four listed cancer types plus the three cancer types in the primary group above.

**Appendix Table 15.** Results of sensitivity analysis that excluded September to November 2021 as a phase-in period. Adjusted differential change in referral rates for regions participating in the trial compared to non-participating regions.

| | Adjusted difference-in-differences, referral rates per 100,000 population (95% CI) | | | | | |
|---|---|---|---|---|---|---|
| **Suspected Cancer Type** | **Months 0 to 5** | **Months 6 to 11** | **Months 12 to 17** | **Months 18 to 23** | **Months 24 to 29** | **Months 30 to 35** |
| **Primary high-detection group** | 20.77 (0.67-40.88, p=0.043) | 14.46 (-7.10-36.03, p=0.177) | 12.29 (-18.19-42.77, p=0.410) | 15.34 (-21.88-52.56, p=0.400) | 20.65 (-22.58-63.88, p=0.331) | 19.34 (-35.38-74.06, p=0.469) |
| Head & neck | 14.75 (0.72-28.79) | 7.36 (-2.50-17.21) | 2.11 (-13.68-17.90) | 7.83 (-9.36-25.03) | 9.81 (-13.39-33.01) | 7.68 (-23.40-38.76) |
| Lung | -0.21 (-5.00-4.59) | 0.17 (-6.70-7.03) | -0.93 (-11.92-10.07) | 1.69 (-13.41-16.79) | 4.36 (-14.05-22.76) | 2.36 (-21.38-26.09) |
| Upper gastrointestinal | 5.29 (-3.92-14.50) | 6.59 (-5.71-18.89) | 11.89 (-3.43-27.21) | 7.83 (-8.78-24.45) | 8.22 (-10.10-26.55) | 9.14 (-10.40-28.67) |
| **Secondary expanded high-detection group*** | 47.06 (-18.00-112.13, p=0.147) | 106.33 (25.28-187.38, p=0.013) | 58.24 (-52.48-168.96, p=0.286) | 90.61 (-25.16-206.38, p=0.118) | 82.02 (-38.55-202.59, p=0.171) | 64.33 (-70.21-198.87, p=0.331) |
| Gynaecological cancer | 4.27 (-8.63-17.17) | 14.97 (-6.97-36.91) | 3.90 (-18.50-26.30) | 16.08 (-10.51-42.67) | 20.43 (-8.25-49.11) | 17.24 (-9.91-44.39) |
| Haematological malignancies (excluding acute leukaemia) | 3.67 (0.01-7.34) | 3.37 (0.29-6.46) | 2.54 (-1.59-6.67) | 4.16 (-0.73-9.06) | 3.34 (-0.78-7.46) | 3.13 (-1.48-7.74) |
| Lower gastrointestinal | 11.34 (-21.75-44.44) | 60.52 (10.92-110.11) | 38.64 (-23.49-100.78) | 33.68 (-21.85-89.21) | 9.87 (-43.67-63.41) | 0.76 (-57.35-58.87) |
| Urological malignancies (excluding testicular) | 8.57 (-3.15-20.28) | 13.63 (-4.24-31.49) | -0.36 (-21.03-20.31) | 19.82 (-1.61-41.25) | 28.05 (0.11-55.99) | 23.21 (-3.50-49.92) |
| **Secondary low-detection group** | 28.68 (-17.63-74.99, p=0.211) | 30.48 (-49.00-109.96, p=0.433) | 27.66 (-49.23-104.55, p=0.462) | 16.60 (-98.27-131.47, p=0.766) | 5.63 (-113.31-124.57, p=0.922) | -9.65 (-136.08-116.77, p=0.875) |
| Acute leukaemia | 0.49 (0.30-0.67) | 0.40 (0.11-0.70) | -0.01 (-0.25-0.23) | -0.09 (-0.62-0.44) | 0.72 (0.14-1.30) | 0.16 (-0.05-0.38) |
| Brain/central nervous system tumours | 0.11 (-2.51-2.74) | 1.42 (-1.52-4.36) | 4.26 (-1.47-9.99) | 0.19 (-3.13-3.51) | 0.25 (-6.56-7.06) | 2.51 (-5.17-10.19) |
| Breast | 1.05 (-18.88-20.97) | 20.51 (-18.46-59.48) | 26.93 (-19.72-73.58) | 19.75 (-24.67-64.18) | 32.08 (-23.71-87.86) | 10.31 (-34.67-55.28) |
| Sarcoma | -0.76 (-8.86-7.35) | -6.65 (-15.65-2.35) | -7.82 (-22.02-6.37) | -15.50 (-36.43-5.42) | -11.39 (-25.35-2.57) | -6.74 (-17.26-3.78) |
| Skin | 28.04 (-14.12-70.20) | 8.58 (-61.58-78.74) | 1.13 (-60.06-62.31) | 1.28 (-99.52-102.07) | -21.77 (-126.11-82.57) | -17.45 (-134.46-99.57) |
| Testicular | 0.38 (-1.43-2.19) | 1.10 (-1.27-3.48) | 0.59 (-1.45-2.64) | 1.77 (-0.88-4.43) | 2.02 (0.20-3.83) | 2.24 (-0.12-4.59) |
| Other | 5.91 (0.59-11.23) | 9.45 (3.30-15.60) | 11.79 (2.59-20.99) | 5.77 (0.04-11.50) | 5.46 (-1.38-12.30) | 4.64 (-0.10-9.38) |

Note: Adjusted differential changes are estimated for the six six-month periods following the start of the trial in month 0. Estimates are calculated at the level of six-month periods (excluding phase-in months) relative to the period prior to the trial start (April 2021 to August 2021, months -6 to -2) using the difference-in-differences model described in methods. The model includes time period and region fixed effects as well as the average

percentage of healthcare staff absent in each region over each time period. Regressions are weighted by population and include clustered standard errors at the regional level.
*The secondary expanded high-detection group consists of the four listed cancer types plus the three cancer types in the primary group above.

**Appendix Table 16.** Results of sensitivity analysis that used the number of hospital beds occupied by patients with confirmed COVID-19 per 100,000 population as an alternate covariate to control for time- and region-varying differences in pandemic-related strain on the healthcare system. Adjusted differential change in referral rates for regions participating in the trial compared to non-participating regions.

| | **Adjusted difference-in-differences, referral rates per 100,000 population (95% CI)** | | | | | |
|---|---|---|---|---|---|---|
| **Suspected Cancer Type** | **Months 0 to 5** | **Months 6 to 11** | **Months 12 to 17** | **Months 18 to 23** | **Months 24 to 29** | **Months 30 to 35** |
| **Primary high-detection group** | 22.74 (-6.43-51.91, p=0.120) | 15.59 (-11.73-42.91, p=0.248) | 13.56 (-17.61-44.73, p=0.375) | 22.19 (-15.08-59.47, p=0.229) | 25.26 (-19.28-69.80, p=0.251) | 20.83 (-33.69-75.35, p=0.435) |
| Head & neck | 13.11 (-2.34-28.57) | 13.31 (-0.56-27.18) | 6.51 (-10.59-23.61) | 12.97 (-3.69-29.63) | 14.15 (-7.15-35.46) | 10.99 (-17.43-39.42) |
| Lung | -0.27 (-6.66-6.13) | 0.81 (-9.62-11.24) | -0.69 (-12.10-10.71) | 3.20 (-11.34-17.74) | 5.62 (-12.98-24.22) | 2.76 (-20.41-25.93) |
| Upper gastrointestinal | 8.75 (-5.09-22.58) | 0.46 (-9.75-10.67) | 7.61 (-6.75-21.97) | 6.40 (-10.31-23.12) | 5.86 (-12.32-24.03) | 5.61 (-15.17-26.38) |
| **Secondary expanded high-detection group*** | 57.40 (-15.29-130.08, p=0.115) | 85.15 (5.18-165.13, p=0.038) | 46.17 (-60.00-152.34, p=0.375) | 83.04 (-37.77-203.85, p=0.167) | 75.24 (-41.14-191.63, p=0.193) | 57.90 (-68.25-184.06, p=0.350) |
| Gynaecological cancer | 6.79 (-9.52-23.09) | 8.56 (-10.20-27.33) | 0.03 (-17.50-17.55) | 14.64 (-8.25-37.54) | 18.88 (-4.23-41.99) | 15.40 (-6.94-37.74) |
| Haematological malignancies (excluding acute leukaemia) | 4.14 (0.98-7.30) | 4.32 (0.64-8.00) | 3.04 (-1.60-7.68) | 4.48 (-0.88-9.85) | 3.61 (-0.80-8.01) | 3.36 (-1.49-8.21) |
| Lower gastrointestinal | 19.81 (-20.84-60.46) | 47.43 (2.81-92.05) | 30.11 (-28.33-88.54) | 25.39 (-33.50-84.28) | 3.31 (-53.28-59.90) | -4.41 (-61.31-52.50) |
| Urological malignancies (excluding testicular) | 7.57 (-3.79-18.94) | 9.48 (-5.46-24.41) | -2.52 (-21.68-16.64) | 16.09 (-8.84-41.03) | 26.28 (-4.52-57.08) | 22.75 (-7.15-52.65) |
| **Secondary low-detection group** | 27.89 (-11.61-67.39, p=0.156) | 23.08 (-50.17-96.33, p=0.518) | 29.65 (-34.30-93.60, p=0.345) | 20.98 (-78.26-120.21, p=0.664) | 11.80 (-90.36-113.96, p=0.812) | -1.45 (-113.02-110.12, p=0.979) |
| Acute leukaemia | 1.16 (0.47-1.85) | 0.06 (-0.16-0.27) | -0.14 (-0.35-0.06) | -0.21 (-0.99-0.58) | 0.27 (-0.53-1.07) | -0.09 (-0.74-0.57) |
| Brain/central nervous system tumours | 1.43 (-0.02-2.89) | 1.96 (-0.18-4.11) | 5.15 (0.16-10.14) | 1.20 (-2.29-4.70) | 1.59 (-4.34-7.53) | 3.68 (-2.77-10.12) |
| Breast | 7.26 (-21.78-36.30) | 12.52 (-23.33-48.37) | 21.11 (-17.76-59.98) | 13.30 (-26.05-52.65) | 26.15 (-22.47-74.77) | 4.94 (-35.45-45.34) |
| Sarcoma | -1.26 (-5.88-3.36) | -0.21 (-4.01-3.58) | -1.19 (-8.99-6.60) | -14.23 (-34.06-5.60) | -9.43 (-20.65-1.79) | -3.96 (-12.33-4.40) |
| Skin | 30.40 (-7.14-67.94) | 8.64 (-56.64-73.91) | 7.30 (-41.87-56.47) | 6.60 (-78.90-92.11) | -15.34 (-106.34-75.66) | -9.10 (-112.60-94.40) |
| Testicular | 0.37 (-1.41-2.15) | 0.58 (-1.85-3.02) | 0.12 (-2.01-2.26) | 1.29 (-1.54-4.12) | 1.57 (-0.29-3.43) | 1.86 (-0.56-4.28) |
| Other | 2.94 (-1.43-7.32) | 6.20 (-1.18-13.57) | 9.24 (-0.16-18.65) | 4.05 (-2.31-10.42) | 4.45 (-2.54-11.45) | 4.48 (-1.35-10.31) |

Note: Adjusted differential changes are estimated for the six six-month periods following the start of the trial in month 0. Estimates are calculated at the level of six-month periods relative to the period prior to the trial start (April 2021 to September 2021, months -6 to -1) using the difference-in-

differences model described in methods. The model includes time period and region fixed effects as well as the number of hospital beds occupied by patients with confirmed COVID-19 per 100,000 population in each region over each time period. Regressions are weighted by population and include clustered standard errors at the regional level.
*The secondary expanded high-detection group consists of the four listed cancer types plus the three cancer types in the primary group above.

**Appendix Table 17.** Results of sensitivity analysis that included no covariate. Adjusted differential change in referral rates for regions participating in the trial compared to non-participating regions.

| | Adjusted difference-in-differences, referral rates per 100,000 population (95% CI) | | | | | |
|---|---|---|---|---|---|---|
| **Suspected Cancer Type** | **Months 0 to 5** | **Months 6 to 11** | **Months 12 to 17** | **Months 18 to 23** | **Months 24 to 29** | **Months 30 to 35** |
| **Primary high-detection group** | 22.40 (-2.24-47.04, p=0.073) | 15.04 (-7.58-37.66, p=0.181) | 13.61 (-17.62-44.85, p=0.374) | 22.59 (-18.62-63.80, p=0.266) | 25.68 (-22.71-74.08, p=0.281) | 21.23 (-37.18-79.65, p=0.457) |
| Head & neck | 12.10 (-2.17-26.37) | 11.90 (0.00-23.81) | 6.95 (-9.44-23.34) | 14.47 (-3.96-32.90) | 15.77 (-7.18-38.73) | 12.60 (-17.56-42.75) |
| Lung | -0.64 (-4.87-3.59) | 0.20 (-6.96-7.36) | -0.70 (-11.70-10.30) | 3.73 (-13.56-21.02) | 6.17 (-14.81-27.16) | 3.27 (-22.24-28.79) |
| Upper gastrointestinal | 9.58 (-4.18-23.35) | 1.84 (-8.78-12.47) | 7.44 (-6.46-21.34) | 5.53 (-10.94-21.99) | 4.91 (-13.60-23.42) | 4.74 (-16.24-25.72) |
| **Secondary expanded high-detection group*** | 62.89 (-5.93-131.71, p=0.071) | 94.38 (15.11-173.65, p=0.022) | 46.01 (-62.85-154.87, p=0.388) | 77.48 (-44.31-199.28, p=0.199) | 69.24 (-49.63-188.11, p=0.239) | 51.93 (-75.67-179.53, p=0.406) |
| Gynaecological cancer | 7.03 (-8.58-22.65) | 11.61 (-8.93-32.14) | 0.56 (-18.85-19.97) | 13.42 (-10.54-37.37) | 17.48 (-6.93-41.89) | 13.97 (-9.06-37.00) |
| Haematological malignancies (excluding acute leukaemia) | 3.94 (0.55-7.32) | 3.89 (0.30-7.49) | 3.05 (-1.68-7.78) | 4.74 (-1.01-10.50) | 3.88 (-0.93-8.70) | 3.63 (-1.55-8.81) |
| Lower gastrointestinal | 22.70 (-18.82-64.23) | 52.29 (7.01-97.58) | 30.04 (-29.09-89.17) | 22.59 (-34.34-79.53) | 0.45 (-55.78-56.68) | -7.45 (-63.02-48.12) |
| Urological malignancies (excluding testicular) | 8.61 (-2.54-19.77) | 11.65 (-3.50-26.81) | -2.50 (-21.58-16.58) | 15.11 (-9.90-40.12) | 25.07 (-6.09-56.24) | 21.65 (-8.37-51.67) |
| **Secondary low-detection group** | 34.27 (-0.83-69.36, p=0.055) | 33.15 (-36.10-102.40, p=0.330) | 29.97 (-36.35-96.29, p=0.357) | 15.93 (-86.50-118.36, p=0.749) | 6.48 (-101.82-114.79, p=0.902) | -7.41 (-122.20-107.38, p=0.894) |
| Acute leukaemia | 1.09 (0.54-1.65) | 0.05 (-0.05-0.15) | -0.29 (-0.58--0.01) | -0.35 (-1.24-0.55) | 0.30 (-0.48-1.08) | -0.21 (-0.83-0.41) |
| Brain/central nervous system tumours | 1.35 (-0.44-3.13) | 1.80 (-0.46-4.06) | 4.70 (-0.02-9.42) | 1.02 (-2.32-4.36) | 1.29 (-4.53-7.10) | 3.41 (-2.90-9.72) |
| Breast | 8.14 (-20.75-37.03) | 14.85 (-22.19-51.89) | 20.86 (-19.04-60.76) | 12.60 (-26.74-51.93) | 25.61 (-23.42-74.63) | 4.56 (-35.79-44.90) |
| Sarcoma | -0.97 (-5.48-3.55) | -0.10 (-4.37-4.17) | -1.14 (-8.81-6.53) | -14.41 (-33.99-5.18) | -9.57 (-20.65-1.52) | -4.09 (-12.50-4.31) |
| Skin | 30.29 (-3.93-64.51) | 13.09 (-46.58-72.76) | 5.44 (-46.06-56.94) | 2.78 (-86.86-92.43) | -18.50 (-114.49-77.50) | -13.23 (-121.13-94.67) |
| Testicular | 0.46 (-1.34-2.26) | 0.71 (-1.83-3.25) | 0.19 (-1.94-2.32) | 1.27 (-1.56-4.09) | 1.55 (-0.32-3.42) | 1.84 (-0.59-4.26) |
| Other | 4.51 (0.33-8.70) | 9.06 (2.31-15.81) | 10.72 (0.35-21.10) | 4.87 (-2.38-12.12) | 5.21 (-2.94-13.36) | 4.86 (-2.48-12.19) |

Note: Adjusted differential changes are estimated for the six six-month periods following the start of the trial in month 0. Estimates are calculated at the level of six-month periods relative to the period prior to the trial start (April 2021 to September 2021, months -6 to -1) using the difference-in-

differences model described in methods. The model includes time period and region fixed effects. Regressions are weighted by population and include clustered standard errors at the regional level.
*The secondary expanded high-detection group consists of the four listed cancer types plus the three cancer types in the primary group above.

**Appendix Table 18.** Results of sensitivity analysis that applied propensity score weights to account for regional imbalances in population size and number of healthcare staff. Adjusted differential change in referral rates for regions participating in the trial compared to non-participating regions.

| | Adjusted difference-in-differences, referral rates per 100,000 population (95% CI) | | | | | |
|---|---|---|---|---|---|---|
| **Suspected Cancer Type** | **Months 0 to 5** | **Months 6 to 11** | **Months 12 to 17** | **Months 18 to 23** | **Months 24 to 29** | **Months 30 to 35** |
| **Primary high-detection group** | 28.65 (1.52-55.78, p=0.040) | 21.04 (5.66-36.43, p=0.010) | 30.35 (3.20-57.49, p=0.030) | 29.66 (-2.97-62.30, p=0.073) | 32.73 (-11.40-76.86, p=0.137) | 33.31 (-15.25-81.86, p=0.168) |
| Head & neck | 14.96 (2.39-27.53) | 13.01 (5.94-20.08) | 13.39 (2.74-24.04) | 19.06 (4.44-33.67) | 20.10 (1.83-38.37) | 19.13 (-5.11-43.36) |
| Lung | 1.28 (-4.73-7.30) | 3.10 (-2.59-8.78) | 2.02 (-7.37-11.41) | 3.61 (-6.13-13.34) | 6.50 (-7.97-20.98) | 6.29 (-9.77-22.35) |
| Upper gastrointestinal | 12.29 (-3.22-27.79) | 4.99 (-8.74-18.72) | 14.90 (-0.83-30.62) | 7.22 (-10.63-25.06) | 6.29 (-16.50-29.08) | 7.99 (-13.37-29.36) |
| **Secondary expanded high-detection group*** | 61.51 (-8.11-131.13, p=0.080) | 107.05 (42.27-171.82, p=0.003) | 81.93 (-8.09-171.95, p=0.072) | 112.65 (11.90-213.40, p=0.030) | 84.83 (-18.54-188.20, p=0.102) | 65.84 (-48.05-179.74, p=0.242) |
| Gynaecological cancer | 4.80 (-12.68-22.29) | 16.73 (-14.52-47.99) | 6.24 (-16.59-29.07) | 15.67 (-10.14-41.48) | 19.23 (-5.03-43.49) | 17.59 (-2.98-38.17) |
| Haematological malignancies (excluding acute leukaemia) | 3.72 (-0.76-8.20) | 4.20 (0.08-8.32) | 3.97 (-1.55-9.50) | 4.96 (-1.35-11.27) | 4.93 (-0.84-10.70) | 5.16 (-1.12-11.43) |
| Lower gastrointestinal | 20.46 (-27.65-68.57) | 47.72 (-11.04-106.48) | 41.12 (-14.49-96.72) | 40.38 (-7.27-88.03) | -2.49 (-50.95-45.96) | -16.04 (-73.60-41.51) |
| Urological malignancies (excluding testicular) | 3.97 (-7.90-15.84) | 17.47 (-2.06-37.01) | 0.31 (-18.97-19.59) | 21.50 (-5.60-48.60) | 29.70 (-5.85-65.25) | 25.47 (-6.26-57.20) |
| **Secondary low-detection group** | 41.80 (10.48-73.12, p=0.011) | 90.15 (-18.99-199.29, p=0.100) | 81.55 (5.79-157.31, p=0.036) | 89.43 (-52.75-231.61, p=0.204) | 61.71 (-55.24-178.67, p=0.284) | 43.36 (-109.07-195.79, p=0.560) |
| Acute leukaemia | 0.37 (-0.06-0.80) | 0.04 (-0.11-0.19) | -0.03 (-0.22-0.17) | 0.21 (-0.33-0.74) | 0.44 (-0.25-1.12) | 0.11 (-0.38-0.60) |
| Brain/central nervous system tumours | 0.71 (-0.46-1.88) | 1.75 (-0.04-3.54) | 2.15 (-1.12-5.42) | 0.33 (-1.74-2.40) | 1.30 (-2.12-4.73) | 1.29 (-3.05-5.63) |
| Breast | 10.42 (-21.26-42.10) | 38.17 (-18.11-94.44) | 43.18 (-5.85-92.22) | 24.75 (-42.54-92.04) | 42.60 (-48.23-133.43) | 11.56 (-60.47-83.60) |
| Sarcoma | -0.08 (-1.96-1.80) | 1.93 (-0.31-4.16) | 1.17 (-3.94-6.28) | 0.73 (-5.26-6.71) | -1.46 (-7.96-5.03) | -2.35 (-10.69-5.99) |
| Skin | 29.86 (3.08-56.64) | 45.15 (-17.75-108.06) | 33.22 (-11.61-78.05) | 64.74 (-27.82-157.29) | 17.88 (-48.54-84.31) | 31.54 (-72.29-135.37) |
| Testicular | 0.50 (-1.35-2.35) | 1.13 (-1.05-3.31) | 0.27 (-1.49-2.02) | 0.65 (-2.26-3.55) | 1.70 (-0.63-4.03) | 1.93 (-1.47-5.33) |
| Other | 1.00 (-0.36-2.36) | 4.43 (0.93-7.93) | 3.81 (-0.96-8.59) | 1.42 (-2.67-5.52) | 2.21 (-0.88-5.29) | 0.56 (-2.89-4.02) |

Note: Adjusted differential changes are estimated for the six six-month periods following the start of the trial in month 0. Estimates are calculated at the level of six-month periods relative to the period prior to the trial start (April 2021 to September 2021, months -6 to -1) using the difference-in-differences model described in methods. The model includes time period and region fixed effects as well as the average percentage of healthcare

staff absent in each region over each time period. Regressions are weighted by propensity score weights and include clustered standard errors at the regional level.
*The secondary expanded high-detection group consists of the four listed cancer types plus the three cancer types in the primary group above.

**Appendix Table 19.** Results of sensitivity analysis that estimated an unweighted regression. Adjusted differential change in referral rates for regions participating in the trial compared to non-participating regions.

| | Adjusted difference-in-differences, referral rates per 100,000 population (95% CI) | | | | | |
|---|---|---|---|---|---|---|
| **Suspected Cancer Type** | **Months 0 to 5** | **Months 6 to 11** | **Months 12 to 17** | **Months 18 to 23** | **Months 24 to 29** | **Months 30 to 35** |
| **Primary high-detection group** | 18.96 (-4.91-42.83, p=0.113) | 12.39 (-6.76-31.54, p=0.192) | 11.50 (-17.94-40.93, p=0.425) | 18.63 (-18.77-56.03, p=0.311) | 28.14 (-13.31-69.59, p=0.172) | 26.65 (-23.76-77.07, p=0.283) |
| Head & neck | 10.88 (-1.25-23.02) | 12.13 (4.00-20.26) | 8.50 (-3.99-21.00) | 14.41 (-0.08-28.91) | 18.56 (1.00-36.12) | 16.65 (-7.86-41.15) |
| Lung | -1.88 (-7.46-3.70) | -0.13 (-6.91-6.65) | -1.82 (-12.63-8.98) | 1.60 (-12.19-15.40) | 2.89 (-14.25-20.03) | 1.96 (-18.48-22.40) |
| Upper gastrointestinal | 9.96 (-4.04-23.97) | 0.47 (-10.27-11.21) | 4.84 (-10.19-19.87) | 2.72 (-14.51-19.96) | 6.74 (-13.78-27.27) | 7.98 (-13.54-29.51) |
| **Secondary expanded high-detection group*** | 51.54 (-23.41-126.49, p=0.167) | 83.51 (3.50-163.52, p=0.042) | 38.23 (-74.46-150.91, p=0.487) | 70.96 (-56.18-198.10, p=0.258) | 73.49 (-52.34-199.32, p=0.237) | 50.81 (-82.49-184.11, p=0.436) |
| Gynaecological cancer | 4.63 (-10.59-19.85) | 8.45 (-10.82-27.73) | -1.13 (-22.73-20.47) | 10.13 (-18.49-38.74) | 16.40 (-10.97-43.76) | 14.99 (-8.64-38.61) |
| Haematological malignancies (excluding acute leukaemia) | 2.65 (0.22-5.09) | 2.76 (0.70-4.82) | 1.76 (-1.43-4.94) | 2.84 (-1.01-6.70) | 2.69 (-0.27-5.66) | 2.16 (-1.11-5.42) |
| Lower gastrointestinal | 15.99 (-29.18-61.16) | 44.17 (-9.24-97.58) | 24.65 (-36.76-86.05) | 21.83 (-35.98-79.64) | -5.68 (-61.03-49.66) | -16.54 (-79.16-46.08) |
| Urological malignancies (excluding testicular) | 9.29 (0.17-18.41) | 15.73 (-3.20-34.66) | 1.47 (-22.22-25.15) | 17.15 (-8.99-43.29) | 31.44 (-4.42-67.29) | 23.28 (-7.71-54.28) |
| **Secondary low-detection group** | 39.21 (3.87-74.54, p=0.031) | 36.19 (-28.46-100.84, p=0.257) | 35.14 (-27.41-97.68, p=0.255) | 26.18 (-72.70-125.06, p=0.587) | 21.41 (-83.36-126.19, p=0.674) | 4.25 (-103.67-112.17, p=0.935) |
| Acute leukaemia | 0.16 (-0.25-0.58) | -0.07 (-0.27-0.13) | -0.15 (-0.46-0.15) | -0.13 (-0.59-0.34) | 0.01 (-0.48-0.50) | -0.22 (-0.62-0.19) |
| Brain/central nervous system tumours | 0.83 (-0.52-2.19) | 1.66 (-0.42-3.74) | 1.93 (-1.53-5.39) | -0.32 (-2.60-1.96) | 0.43 (-3.55-4.42) | 0.54 (-4.19-5.27) |
| Breast | 11.95 (-19.71-43.61) | 8.82 (-27.69-45.33) | 20.28 (-22.79-63.34) | 9.48 (-36.34-55.30) | 25.63 (-31.14-82.40) | 6.33 (-33.02-45.68) |
| Sarcoma | -0.73 (-3.17-1.71) | 0.38 (-2.35-3.12) | 0.99 (-5.27-7.25) | -2.50 (-11.58-6.58) | -2.32 (-9.60-4.97) | -1.24 (-8.68-6.20) |
| Skin | 25.22 (-3.06-53.49) | 20.64 (-33.40-74.68) | 7.45 (-39.03-53.93) | 15.02 (-64.87-94.90) | -7.29 (-93.85-79.26) | -5.87 (-109.71-97.96) |
| Testicular | 0.36 (-1.08-1.81) | 0.53 (-1.65-2.70) | 0.29 (-1.41-1.98) | 0.89 (-1.49-3.27) | 1.91 (0.30-3.51) | 2.62 (0.29-4.95) |
| Other | 1.95 (-0.41-4.31) | 5.53 (0.62-10.44) | 5.30 (-1.27-11.87) | 4.04 (-1.65-9.74) | 3.60 (-2.14-9.35) | 2.35 (-2.69-7.40) |

Note: Adjusted differential changes are estimated for the six six-month periods following the start of the trial in month 0. Estimates are calculated at the level of six-month periods relative to the period prior to the trial start (April 2021 to September 2021, months -6 to -1) using the difference-in-

differences model described in methods. The model includes time period and region fixed effects as well as the average percentage of healthcare staff absent in each region over each time period. Regressions are unweighted and include clustered standard errors at the regional level.
*The secondary expanded high-detection group consists of the four listed cancer types plus the three cancer types in the primary group above.

**Appendix Table 20.** Results of sensitivity analysis that used wild bootstrapping to generate standard errors to account for limited clusters (regions). Adjusted differential change in referral rates for regions participating in the trial compared to non-participating regions.

| | Adjusted difference-in-differences, referral rates per 100,000 population (95% CI) | | | | | |
|---|---|---|---|---|---|---|
| **Suspected Cancer Type** | **Months 0 to 5** | **Months 6 to 11** | **Months 12 to 17** | **Months 18 to 23** | **Months 24 to 29** | **Months 30 to 35** |
| **Primary high-detection group** | 23.71 (-1.63-51.69, p=0.076) | 12.39 (-7.39-29.79, p=0.161) | 10.32 (-17.64-36.58, p=0.435) | 14.46 (-19.17-48.99, p=0.385) | 19.32 (-25.92-66.03, p=0.389) | 17.43 (-31.48-75.96, p=0.516) |
| Head & neck | 12.44 (-4.65-27.90) | 11.05 (0.85-20.15) | 5.83 (-9.20-17.66) | 11.96 (-4.45-27.23) | 13.75 (-7.60-36.47) | 11.41 (-18.93-43.24) |
| Lung | -0.38 (-5.40-4.10) | -0.94 (-8.08-5.61) | -2.02 (-12.72-8.55) | 0.56 (-13.41-14.71) | 3.27 (-14.91-21.88) | 1.26 (-21.24-22.91) |
| Upper gastrointestinal | 10.17 (-3.25-24.79) | 0.96 (-10.19-11.28) | 6.29 (-8.03-20.23) | 2.66 (-16.26-17.36) | 2.85 (-15.51-21.02) | 3.53 (-15.73-26.05) |
| **Secondary expanded high-detection group*** | 62.53 (-22.30-144.27, p=0.124) | 95.01 (11.31-169.19, p=0.032) | 46.83 (-69.71-145.39, p=0.368) | 79.29 (-43.92-193.54, p=0.177) | 70.58 (-55.31-189.56, p=0.225) | 52.76 (-88.43-184.55, p=0.382) |
| Gynaecological cancer | 7.46 (-8.60-24.23) | 11.02 (-11.00-31.63) | -0.10 (-20.02-20.19) | 11.61 (-15.92-38.35) | 16.11 (-7.43-47.04) | 13.14 (-8.99-40.08) |
| Haematological malignancies (excluding acute leukaemia) | 3.98 (0.47-7.71) | 3.80 (0.53-7.60) | 2.94 (-1.91-7.47) | 4.47 (-1.32-10.29) | 3.65 (-0.65-8.95) | 3.49 (-1.08-9.30) |
| Lower gastrointestinal | 21.81 (-24.03-67.00) | 54.62 (6.65-100.48) | 32.83 (-32.32-95.73) | 27.88 (-28.80-84.54) | 4.01 (-51.26-56.39) | -5.29 (-61.35-47.35) |
| Urological malignancies (excluding testicular) | 7.57 (-4.59-19.81) | 13.35 (-3.25-30.62) | -0.65 (-21.72-20.06) | 19.20 (-6.47-43.63) | 27.62 (-1.03-55.32) | 22.94 (-9.09-49.65) |
| **Secondary low-detection group**** | 33.94 (-9.63-69.72, p=0.104) | 33.91 (-37.68-103.56, p=0.320) | 30.97 (-39.00-100.97, p=0.344) | 18.29 (-91.84-120.37, p=0.711) | 8.16 (-122.54-119.38, p=0.874) | -6.45 (-125.72-110.37, p=0.914) |
| Brain/central nervous system tumours | 1.00 (-1.08-4.15) | 1.43 (-1.71-4.52) | 4.26 (-1.68-10.02) | 0.23 (-2.79-2.40) | 0.28 (-6.38-5.97) | 2.49 (-4.97-9.16) |
| Breast | 8.15 (-17.40-38.42) | 14.83 (-23.96-52.30) | 20.84 (-16.40-59.48) | 12.56 (-26.54-53.53) | 25.58 (-21.69-78.82) | 4.54 (-35.45-47.40) |
| Sarcoma | -2.00 (-10.77-6.49) | 0.08 (-4.42-3.91) | -0.97 (-9.94-9.09) | -11.75 (-31.92-9.84) | -7.29 (-18.50-5.74) | -2.29 (-13.91-14.73) |
| Skin | 30.04 (-7.43-67.53) | 14.99 (-48.59-74.29) | 7.61 (-52.62-61.50) | 7.39 (-86.06-104.22) | -15.40 (-123.74-79.92) | -11.11 (-124.84-99.18) |
| Testicular | 0.42 (-1.11-2.60) | 0.76 (-1.61-4.19) | 0.24 (-1.68-2.85) | 1.43 (-1.10-4.77) | 1.66 (-0.19-3.51) | 1.88 (-0.93-4.43) |
| Other | 5.46 (-2.18-9.89) | 10.94 (-0.01-18.82) | 12.88 (-1.45-24.33) | 6.52 (-4.01-13.24) | 6.26 (-5.24-16.12) | 5.38 (-4.16-11.78) |

Note: Adjusted differential changes are estimated for the six six-month periods following the start of the trial in month 0. Estimates are calculated at the level of six-month periods relative to the period prior to the trial start (April 2021 to September 2021, months -6 to -1) using the difference-in-differences model described in methods. The model includes time period and region fixed effects as well as the average percentage of healthcare staff absent in each region over each time period. Regressions are weighted by population and use wild bootstrapping to generate standard errors to account for limited clusters (regions).

*The secondary expanded high-detection group consists of the four listed cancer types plus the three cancer types in the primary group above.
**We were unable to produce results of wild bootstrap analysis for individual analysis of acute leukaemia, which had the smallest number of referrals of all individual cancer types.

**Appendix Methods 3.** Back-of-envelope calculation of expected number of new referrals due to positive Galleri tests in the first year of the trial for the purposes of plausibility checks.

Number of participants in intervention arm: 70,000[1]

Positivity rate of Galleri test: 1.4%[8]

Referrals following positive Galleri test (Num participants * Positivity Rate): ~980

Assumed proportion with suspected cancer site in primary high-detection group: ~10-40%

Referrals for primary high-detection cancer types following positive Galleri test: ~100-400

Note: There is limited public information available to estimate what proportion of positive Galleri tests in a non-symptomatic screening population are for suspected cancer sites in the primary high-detection group. The assumed proportion used in this calculation encompasses a wide range that is roughly centered on the proportion of true positive Galleri tests resulting in new cancer diagnoses that were among the primary high-detection cancer types (7 out of 29, or 24%) reported in the PATHFINDER I study.[8]

**Appendix Methods 4.** Changes made to analysis plan when compared to preregistration and prior analysis.

Our study ("Potential spillover effects on diagnostic delay for cancer during the NHS-Galleri trial: a quasi-experimental difference-in-differences study") follows an analysis plan that was registered on the Open Science Foundation (OSF) website on August 30, 2025.[9]

The following changes were made to our study which were not reflected in the preregistered analysis plan:

First, in the analysis reported in our study, we excluded referrals in which the suspected cancer type field contained the following values: "Missing or Invalid", "Suspected cancer – non-specific symptoms", and "Suspected children's cancer". This change had no effect on our analyses of our primary high-detection group of cancer types, our secondary expanded high-detection group of cancer types, or our secondary low-detection group of cancer types, as these groups only included referrals for suspected cancer associated with a specific tumor site. In practice, this change only meant that we did not conduct exploratory analyses of referrals for these missing/invalid or non-site specific categories of suspected cancer.

Second, in the analysis reported in our study, we included one additional sensitivity analysis as an extra robustness check. In this additional sensitivity analysis, we used unweighted regressions, in contrast to the primary analysis which used regressions weighted by referral volume (for analysis of diagnostic delay rates) or regional population (for analysis of referral rates).

Prior to registering the analysis plan for the current study, three study team members (Sean Mann, Beth Ann Griffin, and Joshua Eagan) conducted a preliminary analysis that was highly similar to the analysis described in this study. The results of this preliminary analysis were made publicly available as a preprint on November 7, 2024.[10] Summary results of this preliminary analysis were also presented by Sean Mann in a conference presentation at the AcademyHealth annual meeting in July 2024[11] and in a NIH Pragmatic Trials Collaboratory grand rounds talk in March 2025.[12]

That preliminary analysis features the same dataset, primary outcome, covariate, and event study difference-in-differences analytic design that is described here. There are four main differences between the preliminary analysis and the analysis described in this study:

First, the preliminary analysis was conducted on a dataset that included a period of 2.5 years (April 2021 – September 2023), in contrast to the 3.5 year period used in our study (April 2021 to September 2024). The period used in our preliminary analysis and the period used in our current study reflected the full period for which final wait times data were available at the time of each analysis.

Secondly, the preliminary analysis focused on the primary stratified group of less commonly referred cancers that featured lower referral volumes (Gynaecological, head and neck, urological, upper gastrointestinal, lung, hematological, sarcoma, brain/nervous system, children's cancer, testicular, acute leukemia, and cancers in the 'other' category) rather than the primary high-detection group of cancer types used in this study (head and neck, lung, upper gastrointestinal). The preprint describes the stratification approach used in the preliminary analysis as follows:

"We examined three primary outcomes in our analyses: diagnostic delay rates for all cancers, for the 'most commonly referred' cancers, and for all other 'less commonly referred' cancers. Our concern over potential spillover from the NHS-Galleri trial was particularly focused on types of suspected cancers detected by the Galleri test for which there was limited existing diagnostic capacity. In the absence of publicly available data on diagnostic capacity by type of suspected cancer, we relied on the volume of referrals that resulted in diagnostic resolution as an indicator of capacity. Referral volume was then used to stratify referrals into two groups according to whether they were for one of the 'most commonly referred' cancers or for any other 'less commonly referred' cancer."

The results of this preliminary analysis, as summarized in the preprint abstract, are reproduced as follows: "We observed no significant differences in rates of diagnostic delay for all cancers or for the most commonly referred cancers. For less commonly referred cancers, the percentage of patients experiencing diagnostic delays increased from 32.6% to 35.1% in regions participating in the NHS-Galleri trial from the 6 months before to the 6 months after trial start, compared with a small reduction from 32.7% to 31.7% in regions that did not participate (adjusted difference 3.4 percentage points, 95% confidence interval 1.2 to 5.7). These differences persisted from 6 to 12 months after trial start and receded the following year."

In our current study, we changed our stratification approach to focus on the prespecified cancers from the trial protocol,[1] which were identified by the Galleri test developers as "having higher cancer signal detection" for cfDNA-based MCED testing.[13] This was done to address potential concerns that the focus on less commonly referred cancers was an arbitrary choice. By making this change, we let the NHS-Galleri trial investigators decision to focus their stratified primary analysis on a select group of high-detection cancers guide our own analytic design. Our current focus on the cancers from the trial protocol also follows the approach of another recent independent analysis of the NHS-Galleri trial.[14]

We further focused our study analysis on a primary analytic grouping that consisted of cancer types that exclusively contained cancers from the trial protocol and were not subject to routine single-cancer screening. We also conducted analyses of a secondary expanded grouping of all cancer types that included cancers from the trial protocol, even if these cancer types also included cancers not in the protocol or were subject to routine single-cancer screening. A final set of stratified analyses reported results for a secondary group of low-detection cancer types that did not contain any cancers from the trial protocol.

Third, we included analysis of a secondary outcome (referral volume), to help understand whether changes in referral volume were a potential driver of any observed changes in our primary outcome (diagnostic delay rates). Our primary outcome of interest, diagnostic delay rates, remained the same.

Fourth, we included several additional sensitivity analyses to help determine the robustness of findings from our primary analysis. This included additional sensitivity analyses that: used the number of hospital beds occupied by patients with confirmed COVID-19 per 100,000 population as an alternate covariate to control for time- and region-varying differences in pandemic-related strain on the healthcare system; applied propensity score weights to account for regional imbalances in population size and number of healthcare staff; estimated an unweighted regression; used wild bootstrapping to generate standard errors to account for limited clusters (regions); used the alternative 14-, 42-, and 62-day cutoffs to calculate diagnostic delay rates; and used average waiting times calculated by mid-point imputation as an alternative outcome specification (appendix methods 2). We also retain the two sensitivity analyses from our preliminary analysis, including models that: excluded September to November 2021 as a phase-in period; used no covariate.
